# Predicting coronary artery disease severity through genomic profiling and machine learning modelling: The GEnetic SYNTAX Score (GESS) trial

**DOI:** 10.1101/2024.12.04.24318505

**Authors:** Fani Chatzopoulou, Nikolaos Mittas, Dimitrios Chatzidimitriou, Alexandros Giannopoulos-Dimitriou, Aikaterini Saiti, Maria Ganopoulou, Efstratios Karagiannidis, Andreas S. Papazoglou, Nikolaos Stalikas, Anna Papa, George Giannakoulas, Georgios Sianos, Lefteris Angelis, Ioannis S. Vizirianakis

**Affiliations:** Laboratory of Microbiology, School of Medicine, Aristotle University of Thessaloniki, Thessaloniki, Greece; Labnet Laboratories, Thessaloniki, Greece; Hephaestus Laboratory, School of Chemistry, Faculty of Sciences, Democritus University of Thrace, Kavala, Greece; Laboratory of Pharmacology, School of Pharmacy, Aristotle University of Thessaloniki, Thessaloniki, Greece; School of Informatics, Aristotle University of Thessaloniki, Thessaloniki, Greece; Second Department of Cardiology, Hippokration General Hospital, Aristotle University of Thessaloniki, Thessaloniki, Greece; AHEPA University General Hospital of Thessaloniki, 1st Cardiology Department, Thessaloniki, Greece; Department of Health Sciences, School of Life and Health Sciences, University of Nicosia, Nicosia, Cyprus

**Keywords:** Coronary artery disease, CAD, SYNTAX score, SNPs, biomarkers, cardiovascular disorders, CVDs, risk prediction, machine learning, personalized (precision) medicine, pharmacogenomics

## Abstract

Cardiovascular diseases (CVDs) present multifactorial pathophysiology and produce immense health and economic burdens globally. The most common type, coronary artery disease (CAD), shows a complex etiology with multiple genetic variants to interplay with various clinical features and demographic traits affecting CAD risk and severity. The development and clinical validation of machine learning (ML) algorithms that integrate genetic biomarkers and clinical features can improve diagnostic accuracy for CAD avoiding, thereby, unnecessary invasive procedures. To this end, we present, here, the development of a data-driven ML approach able to predict the existence and severity of CAD based on the analysis of 228 single nucleotide polymorphisms (SNPs) and clinical and demographic data of 953 patients enrolled in the Genetic Syntax Score (GESS) trial (NCT03150680). Two competing ensemble models (one with clinical predictors and another with clinical plus genetic predictors) were built and evaluated to infer their prediction capabilities. The ensemble model with both clinical and genetic predictors exhibited superior diagnostic performance compared to the competing model with only clinical predictors. The proposed ML framework identified a total of eight contributing SNPs as predictors for the existence of obstructive CAD and seven significant SNPs for the severity of CAD. Such algorithms positively contributes to global efforts aiming to predict the risk and severity of CAD in early stages, thus lowering the cost as well as achieving prognostic, diagnostic, and therapeutic benefits in healthcare and improving patient outcomes in a non-invasive way. Overall, the design and execution of this trial reinforces clinical decision-making and facilitate the harmonization in digitized healthcare within the concept of precision medicine.

**Clinical Trial Registration:** NCT03150680; https://clinicaltrials.gov/study/NCT03150680?cond=NCT03150680&rank=1

## Introduction

Several machine learning (ML) approaches have already been applied in the clinical setting, including cardiovascular disorders (CVDs). In particular, the utility of ML methodologies has been shown in imaging practices used for the diagnosis of acute coronary syndrome (ACS) and in the prediction of coronary artery disease (CAD) severity. Indeed, emerging evidence suggests that ML methodologies may accurately predict CAD severity as well as short- and long-term outcomes in patients presenting with ACS^1–5^. Specifically, ML algorithms have been applied in the CAD setting to: (i) predict the occurrence of obstructive CAD by evaluating multiple clinical variables and the coronary artery calcium score^6^; (ii) improve functional coronary assessment, as well as to detect lesion-specific ischemia by using computational fluid dynamics algorithms^7,8^; (iii) estimate the pre-test probability of CAD^9^; and (iv) evaluate the automatic prediction of obstructive CAD from myocardial perfusion imaging^10,11^.

A previous effort by our research group led to the development of a ML clinical risk-stratification framework aiming to the prediction of the severity of CAD based on the assessment of SYNTAX score^12–15^. By combining anatomic and clinical prognostic variables, the SYNTAX score is used to predict the prognostic course in patients with CAD and guide practitioners in choosing percutaneous coronary intervention (PCI) or coronary artery bypass graft (CABG) surgery. Higher scores are associated with more complex CAD^13^. At this end, we proposed an ensemble ML approach that integrates via a two-stage model both classification and regression techniques into a unified risk-score assessment process in order to model: (i) the existence of obstructive CAD through a binary classifier (patients with zero vs. non-zero SYNTAX score); and (ii) the severity of CAD (the expected SYNTAX score) through a regression model given that a patient is diagnosed with the existence of obstructive CAD (non-zero SYNTAX score).

The rationale behind this work was to further extend our previous research efforts through the leverage of genomic information hidden into single nucleotide polymorphisms (SNPs) analyzed using next generation sequencing (NGS) technology. The aim of the current study was to investigate whether introducing the genetic information of specific SNPs into the development of ML prediction models [i.e., including clinical biomarkers and demographic parameters (Supplementary Table 1)] could bolster SYNTAX score prediction in relatively large patient populations. This effort was achieved by evaluating 228 SNPs (Supplementary Table 2, previously recognized as contributing factors to CAD from genome-wide association studies (GWAS), in 953 patients enrolled in the GESS trial (NCT03150680)^13^. The findings obtained contribute to non-invasive evaluation of CAD severity through ML approaches that allows practitioners very early to carry out personalized intervention in the clinical setting, stratification, and therapeutic guidance of their patients. It also provides a way in which the integration of genetic data of SNPs into ML models, may reinforce clinical decision-making and facilitate the harmonization in digitized healthcare^16^ in compliance with the American Heart Association recent statement on AI/ML use in healthcare^17^.

## Methods and Materials

### Clinical study design and data collection

The detailed design of the GESS trial (ClinicalTrials.gov Identifier: NCT03150680) has been previously published^13^. Adult patients undergoing invasive coronary angiography were enrolled in this trial after providing written informed consent. Two experienced interventionalists blinded to the study protocol assessed the obtained angiographic images, and, thereby, the SYNTAX score was calculated for every study participant. Patients with history of prior revascularization procedure and patients with cardiopulmonary arrest on admission were not deemed eligible for enrolment. Ethical approval was obtained from the Scientific Committee of the AHEPA University Hospital of Thessaloniki, Greece (reference number 309/11–05-2017); all trial procedures complied with the Declaration of Helsinki^18^.

Pre-specified clinical data, including demographic characteristics, medical history, clinical presentation (Supplementary Table 1) and medication, were recorded for the entire study population under standardized methods. Additionally, peripheral blood samples were drawn on the enrolment day -prior to coronary angiography-for genomic profiling. The vials of drawn blood were aliquoted and stored as whole blood, plasma, serum, and buffy coat, accordingly.

### NGS analysis of SNP variations in patient DNA samples

Genomic DNA was extracted from blood using the QIAamp DNA Blood Midi kit (Qiagen) following the manufacturer’s instructions. The extracted samples were quantified using Nanodrop 1000 (Thermo Scientific).

### Library preparation

A custom-made panel consisting of 228 SNPs was used (Supplementary Table 2). SNP selection was based on their potential correlation to the pathophysiology of CVDs, as previously presented^13,14^. DNA libraries were built using the QIAseq Targeted DNA Panel kit (Qiagen) following the manufacturer’s protocol. In brief, DNA (40 ng) was enzymatically fragmented, end repaired and A-tailed followed by adapter ligation. These adapters (12 different indices, IL-N701–N715) are unique molecular indices (UMIs) that integrate on each samples sequence. Then, double cleanup was performed using QIAseq beads and freshly prepared ethanol. Following the cleanup, target enrichment was performed using one region-specific primer and one universal primer complementary to the adapter. After a cleanup step, a universal PCR was carried out to amplify the library and add index primer (IL-S502–S511). The reaction was performed directly in the QIAseq 96-index I set A plate, that contains sample index primer and universal PCR primers. After completion of universal PCR, a final cleanup step using QIAseq beads was performed.

### Sequencing and data analysis

Amplified libraries were quantified using the Qubit dsDNA HS (High Sensitivity) assay kit and the Qubit 2.0 Fluorometer (Thermo Fisher Scientific). In addition to fluorometry, libraries were further quantified by qPCR, using the Universal qPCR Master Mix (KAPA Library Quantification Kit) for Illumina Platforms in two working dilutions. The libraries were normalized, pooled, and diluted to 1 nM. After denaturation with 0.1N NaOH the pool was further diluted to 1.3 pM. The library was loaded onto a Mid Output cartridge on the MiniSeq system (Illumina) according to manufacturer’s instructions and sequenced using paired-ends (2⍰×⍰151 bp) and the QIAseq custom primer Read 1 provided with the QIAseq library kit. Upon completion of the sequencing run, FASTQ files were downloaded for further bioinformatic analysis. NGS data (FASTQ files) were analyzed using the CLC Genomics Workbench v.20.0.3 (Qiagen) with the Biomedical Genomics Analysis Plugin v. 20.0.1. A custom workflow has been created for trimming and annotation of UMIs, mapping to hg19 (reference genome), removal of ligation artefacts, detection, and annotation of indels and structural variants, and local realignment. The identified variants were called, and the three different genotypes (homozygous for the reference allele, heterozygous, homozygous for the alteration) were recognized.

### Machine learning framework

The data-driven ML framework along with its core elements aiming at the prediction of SYNTAX score is illustrated in Fig. 1. The methodology follows previous efforts of our group^12,15^, involving nine phases with important actions and decisions that should be cautiously considered to design and implement a ML solution with the aim of efficiently achieving the goal of CAD risk and severity prediction at real time and non-invasively in the point-of-care. Following the structured process for the design, development and deployment of any ML solution, the workflow of the proposed approach can be summarized into the following nine phases: (i) Problem Definition, (ii) Data Collection, (iii) Data Preparation, (iv) Data Exploration, (v) Model Selection, (vi) Feature Selection (vii) Model Building, (viii) Model Evaluation and (ix) Practical Implications of model deployment (Fig. 1). Note that the detailed description of the ML workflow along with its phases and decisions made for each phase are presented in the Supplementary data section (Supplementary note). In brief, the proposed ML solution adopts the idea of ensemble learning, where multiple models (base learners) co-operate with the aim of achieving superior performance compared to any individual model. In our framework, the ensemble mechanism exploits (a) a classification base learner (or zero-part model) that is responsible for discriminating patients into two mutually exclusive groups (patients with SYNTAX score higher than zero versus patients with SYNTAX score equals to zero) and (b) a regression base learner (count-part model) that is dedicated to the prediction of the expected SYNTAX score given that a patient is classified into the non-zero group (SYNTAX score higher than zero).

**Figure 1.**
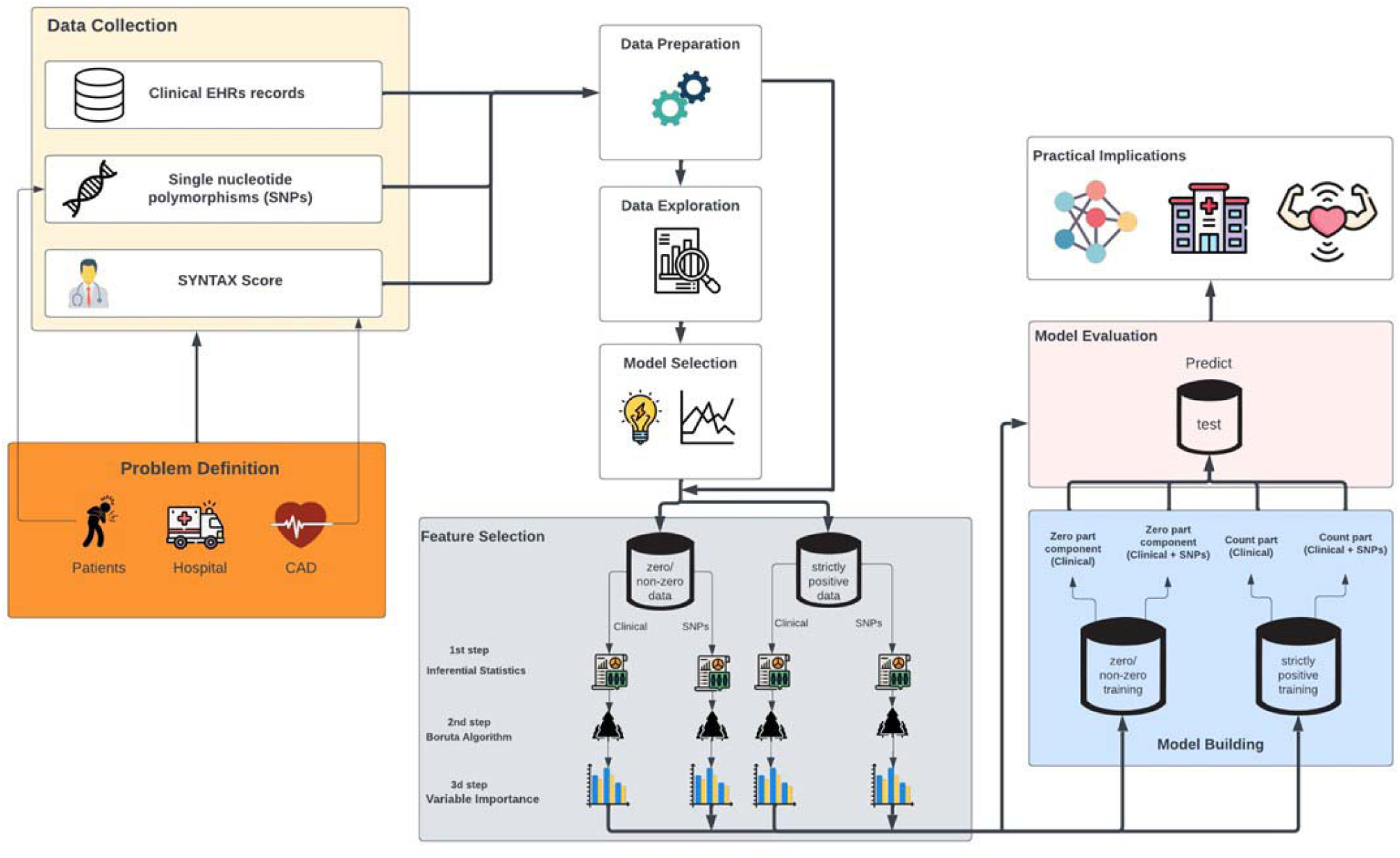
Data-driven ML approach of the study.

## Results

### ML model functionality and practical utility

In this section, we mainly focus on the presentation of the results concerning the identification of the set of the important clinical- and genome-related factors (Feature Selection phase) that were used, in turn, as inputs into the fitting of the competing ensemble models (Model Building). Supplementary Fig. S1 presents the distribution of SYNTAX score for the total number of patients participated in the current study (red violin plot) compared to the SYNTAX score distribution for the subgroup of patients that presenting SYNTAX score higher than zero (blue violin plot). Also, Supplementary Table 3 summarizes the descriptive statistics for the selected clinical and demographic variables from various types of Electronic Health Records (EHRs).

Supplementary Tables 4 and 5 show the initial set of candidate risk-factors that exhibited a statistically significant effect on the SYNTAX score response variables for the zero- and count-part models, respectively. In the case of the zero-part model (Supplementary Table 4), the Likelihood Ratio (LR) test after the fitting of separate univariate logistic models for each candidate predictor indicated that a set of forty-one predictors (nineteen clinical risk-factors and twenty-two SNPs) seem to meet the first criterion for insertion into the second round of the feature selection protocol. Concerning the count-part model, thirty-three predictors (Supplementary Table 5) (twelve clinical risk-factors and twenty-one SNPs) were identified as significant risk-factors for further investigation into the second round of the feature selection protocol. At the second step, the Boruta algorithm was executed on the candidate sets of risk-factors that were extracted from the previous selection round to decide upon the final predictors that were inserted into the zero- and count-part models of the ensemble mechanism. The algorithm resulted into the variable importance measure (VIM) value for each predictor assessing the contribution of features to the modeling process of the training phase^19^. More specifically, the wrapper feature selection technique was applied, separately, for the subsets of clinical risk-factors and SNPs after omitting clinical predictors that either (a) presented a high proportion of missing observations that would lead, in turn, to well-known problems related to the fitting of multivariate models or (b) participated into the computation formulae of RATIO1 (monocyte to HDL cholesterol ratio), RATIO2 (lymphocyte to monocyte ratio), RATIO3 [atherogenic index of plasma levels, *log*(*TG / HDL*)] and RATIO4 [liver cell injury indicator, serum aspartate aminotransaminase (SGOT) to serum alanine aminotransaminase (SGPT) ratio], so as to mitigate the arisen multicollinearity issues, due to the inclusion of intercorrelated features.

The results related to the distributions of VIM after the execution of the Boruta algorithm on the qualified sets of the clinical and SNPs predictors for the zero- and count-part models are graphically displayed in Fig. 2a. From the total set of the nineteen clinical predictors, fourteen were confirmed as statistically significant for insertion into the zero-part of the model (upper left, green-colored boxplots in Fig. 2a). Additionally, the Boruta algorithm signified eight important SNPs (upper right, green-colored boxplots in Fig. 2a) that will be used for the model building phase of the zero-part model. Regarding the regression base learner (count-part model), nine out of twelve clinical risk factors (lower left, green-colored boxplots in Fig. 2a) and seven out of twenty-one SNPs (lower right, green-colored boxplots in Fig. 2a) were confirmed as informative.

**Figure 2a.**
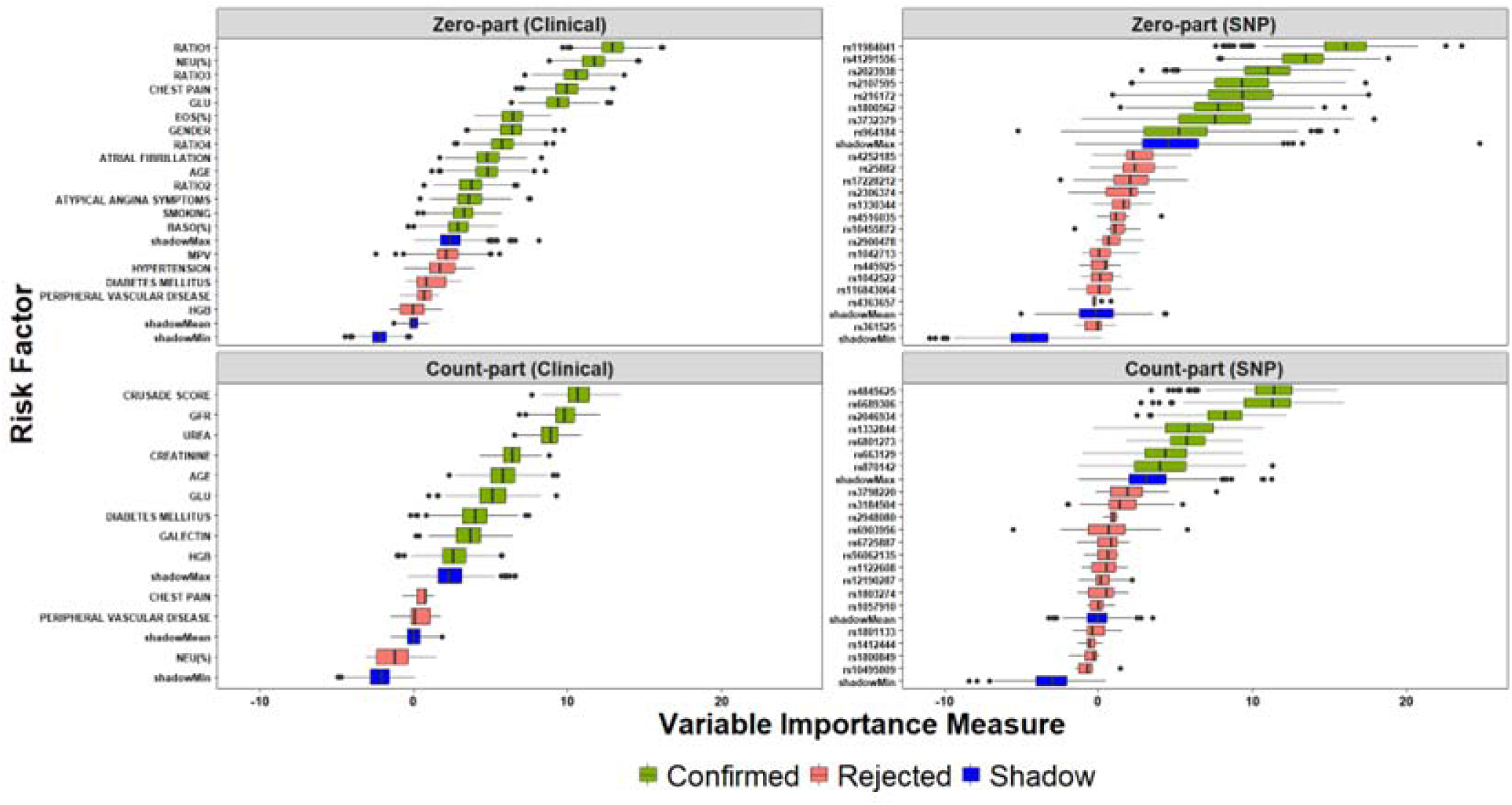
Distributions of variable importance measure for the set of Clinical and SNPs predictors fo the zero- and count-part models.

The above sets of important clinical risk-factors and SNPs constituted the basis for the training of the two competing ensemble models, called Model A (ensemble model with clinical predictors) and Model B (ensemble model with clinical plus SNPs predictors). The examination of the performance metrics for the classification base learner (zero-part model) (Supplementary Table 6) indicates superior predictive capabilities for Model B that exploited both clinical and SNPs information for all performance indicators compared to Model A. The graphical inspection via receiver operating characteristics (ROC) analysis (Fig. 2b, left figure) that visualizes the trade-off between true positive rate (TPR) (or sensitivity) and false positive rate (FPR) (or 1– specificity) suggests that both classifiers present sufficient discrimination capacity compared to the baseline random classifier (points lying along the diagonal reference line, where FPR = TPR), whereas Model B seems to dominate Model A, since it is closer to the optimal top-left corner of the plot. Indeed, the computation of the area under the curve (AUC) measure for the two competing classifiers confirms that Model B presents a higher AUC value (*AUCROC*_Model B_ = *0.798* ) compared to the corresponding value derived from the model incorporating only the set of clinical risk-factors (*AUCROC*_Model A_ 0.757), a difference that was statistically signified via the execution of the Delong’s test (*Z* = 2.451, *p* = 0.014). Moreover, the investigation of the performance capabilities of the two classifiers through precision-recall (PR) curves (Fig. 2b, right figure) showcases similar findings, since Model B is closer to the top-right corner presenting, again, higher AUC value (*AUCPR*_Model B_ = 0.912) in comparison with Model A (*AUCPR*_Model A_ = 0.893). We note that in the case of PR curves, the baseline model (horizontal reference line) represents the performance of a model classifying all instances to the positive class.

**Figure 2b.**
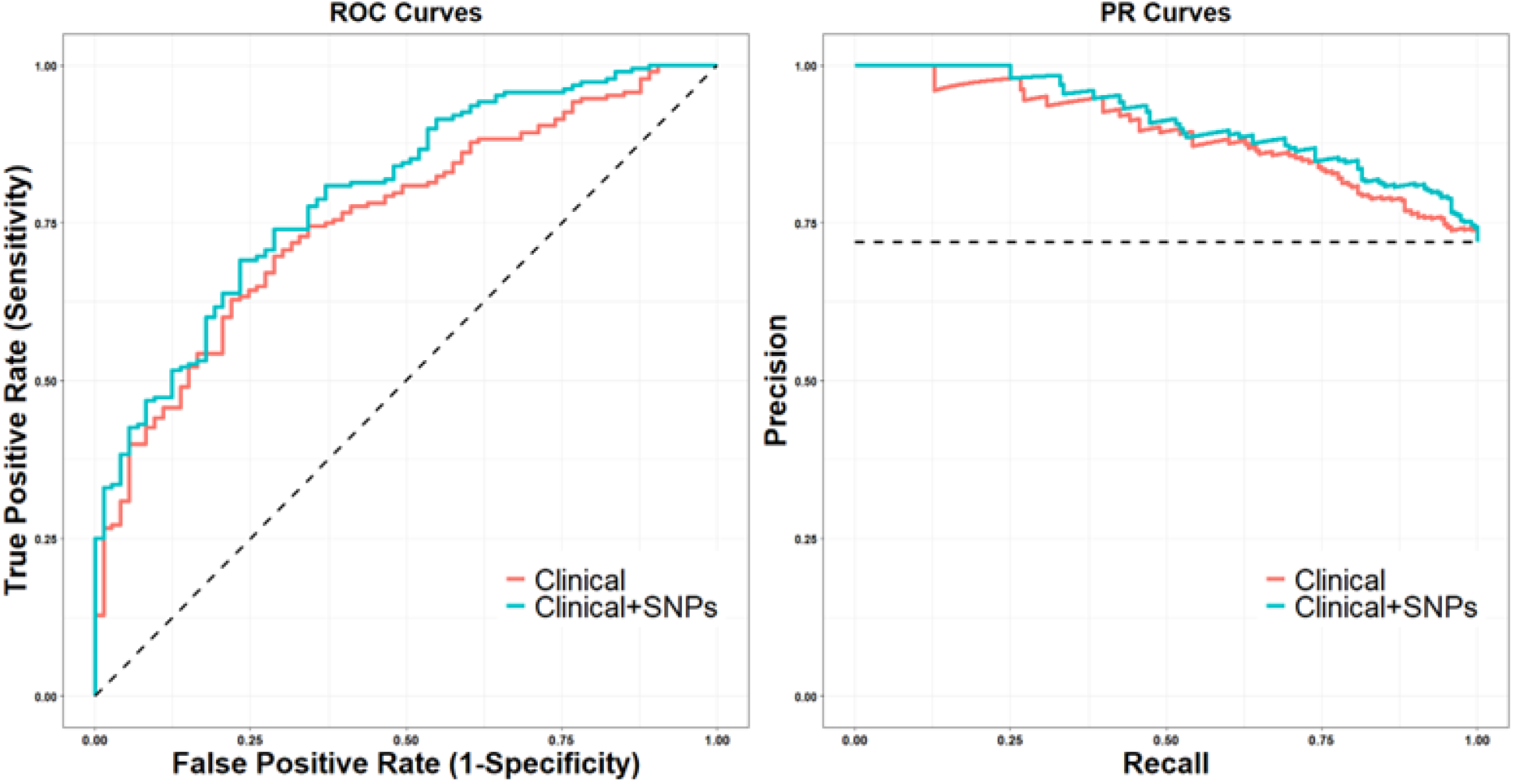
ROC and PR curves for the evaluation of the classifier (zero-part model) for the competing models (Clinical (Model A)/Clinical+SNPs (Model B)).

The performance evaluation concerning the competing regression base learners that are responsible for the prediction of the expected SYNTAX score for patients with obstructive CAD revealed, again, that Model B yielded superior prediction performance compared to Model A in terms of both MdAE and MdMRE values (Supplementary Table 6). The conduction of appropriate inferential mechanisms via the Wilcoxon Signed Rank test demonstrated statistically significant differences for the distributions of both the absolute error (*V* = 9696, *p* = 0.008) and the magnitude of relative error (*V* = 10095, *p* < 0.001). Finally, the comparison related to the prediction capabilities of the ensemble learning synergy between the classification and regression base learners showed, again, that the model with both clinical and SNPs predictors (Model B) outperformed Model A that leveraged only clinical risk-factors (Model A) (*V* = 16731, *p* = 0.002).

### Network analysis of the important SNPs predictors of the zero- and count-part models

The genetic etiology of CAD includes multiple genetic variants associated with CAD risk and severity^20–23^. The eight and seven important SNPs (Table 1) that were inserted into the zero- and count-part models, respectively, were subjected to bioinformatics analysis to determine their corresponding genes accompanied by their genetic/protein interactors, as well as to unveil the association between a variant with human diseases and drug phenotype.

**Table 1.** Genomic characteristics of the important SNP predictors in the ML risk-stratification model.

| Model | SNP | Gene (within/nearby) | Gene name | Position (GRCh38) | Consequence | MAF* |
| --- | --- | --- | --- | --- | --- | --- |
| Zero-part | rs3732379 | CX3CR1 | C-X3-C Motif Chemokine Receptor 1 | chr3:39265765 | missense variant | 0.14 (T) |
|  | rs1800562 | HFE | Homeostatic Iron Regulator | chr6:26092913 | missense variant | 0.01 (A) |
|  | rs11984041 | HDAC9 | Histone Deacetylase 9 | chr7:18992312 | intron variant | 0.10 (T) |
|  | rs2023938 | HDAC9 | Histone Deacetylase 9 | chr7:18997152 | 3' prime UTR variant | 0.11 (C) |
|  | rs2107595 | Mapped genes: HDAC9, TWIST1 | Histone Deacetylase 9, Twist family bHLH transcription factor 1 | chr7:19009765 | regulatory region variant | 0.25 (A) |
|  | rs41291556 | CYP2C19 | Cytochrome P450 Family 2 Subfamily C Member 19 | chr10:94775416 | missense variant | < 0.01 (C) |
|  | rs964184 | ZPR1 | ZPR1 Zinc Finger | chr11:116778201 | 3' prime UTR variant | 0.22 (G) |
|  | rs216172 | SMG6 | SMG6 Nonsense Mediated MRNA Decay Factor | chr17:2223210 | intron variant | 0.34 (C) |
| Count-part | rs6689306 | IL6R | Interleukin 6 Receptor | chr1:15442347 | intron variant | 0.42 (A) |
|  | rs4845625 | IL6R | Interleukin 6 Receptor | chr1:154449591 | intron variant | 0.44 (T) |
|  | rs2046934 | MED12L - P2RY12 | Mediator Complex Subunit 12L - Purinergic Receptor P2Y12 | chr3:151339854 | intron variant | 0.13 (G) |
|  | rs6801273 | MED12L - P2RY12 | Mediator Complex Subunit 12L - Purinergic Receptor P2Y13 | chr3:151345042 | intron variant | 0.42 (C) |
|  | rs870142 | STX18-AS1 | STX18 Antisense RNA 1 | chr4:4646320 | intron variant | 0.20 (T) |
|  | rs1332844 | PHACTR1 | Phosphatase and Actin Regulator 1 | chr6:12888772 | intron variant | 0.32 (C) |
|  | rs663129 | Mapped genes: RNU4-17P, MC4R | RNA U4 small nuclear 17 pseudogene, Melanocortin 4 receptor | chr18:60171168 | intergenic variant | 0.25 (A) |

\* MAF, minor allele frequency (minor allele)

Regarding the zero-part model (classifier), three out of eight SNPs are located either within the intronic (rs11984041) or the 3’ prime untranslated (3’-UTR) (rs2023938) or the intergenic region (rs2107595) of the *HDAC9* gene. The genetic variant rs2107595, localized between the *HDAC9* and *TWIST1* genes, is a known risk factor associated with CAD, large artery intracranial occlusive disease^24^, and aortic calcification^25,26^. Different studies, however, contradict whether this SNP regulates the *HDAC9* or the *TWIST1* mRNA expression^26^. In addition, rs3732379, rs1800562, and rs41291556 are nonsynonymous variants of the genes *CX3CR1*, *HFE*, and *CYP2C19*, respectively, causing alterations in the amino acid sequence of the corresponding proteins. Besides, the variant rs964184 is localized in the 3’-UTR of the ZPR1 gene and the rs216172 variant inside the intronic region of the *SMG6* gene.

In the count-part model (regressor), six out of seven important SNPs are intronic. Specifically, (i) the variants rs4845625 and rs6689306 are localized in the *IL6R* gene; (ii) the variants rs2046934 and rs6801273 are localized in the overlapping genes (OLGs) *MED12L* and *P2RY12*; (iii) the variant rs870142 is located in the interval between *STX18* and *MSX1* and, according to a previous report, potentially affects the expression of the latter^27^; and (iv) the variant rs1332844 which is located in the *PHACTR1* gene. Finally, rs663129 is an intergenic SNP located downstream of *PMAIP1* and *MC4R* genes.

According to the network analysis, based on DisGeNET database^28^, the important SNP predictors of the zero-part model are mainly associated with CVDs, including CAD and atrial fibrillation (Fig. 3a). However, compared to the variant-disease network of the zero-part model, the SNP predictors of the count-part model participate in a more compact variant-disease network which highlights the strong association of these variants particularly with CVDs (Fig. 4a). The expert-curated interactions of the genes corresponding to the important SNPs predictors of the zero- and count-part models were retrieved from BioGRID database of protein, genetic and chemical interactions^29^ and integrated into graphical network representations (Fig. 3b and Fig. 4b, respectively).

**Figure 3.**
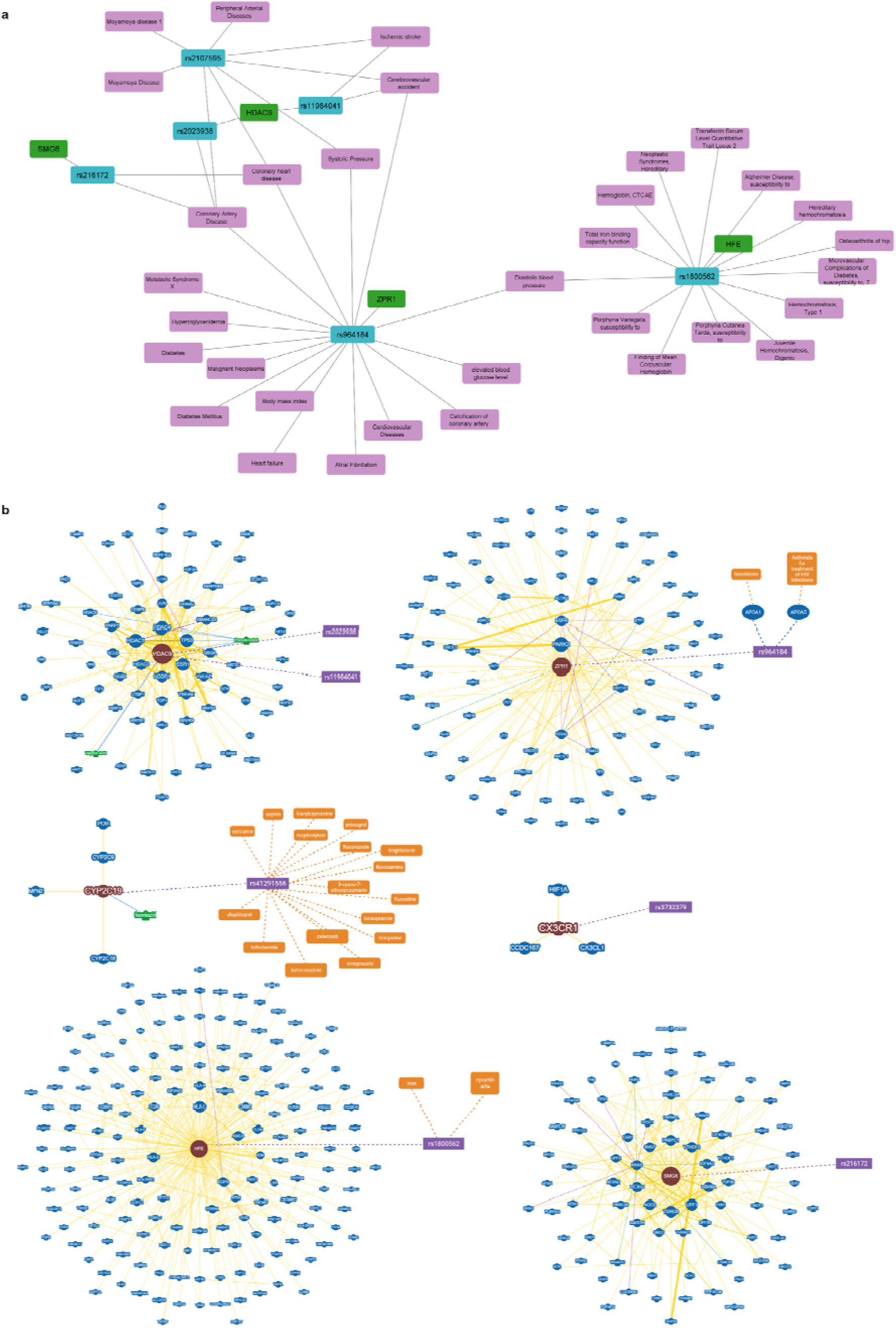
(a) Network visualization of the zero-part SNPs and their association with diseases according to DisGeNET database. The disease annotations, the SNPs, and the genes corresponding to the SNPs are depicted in purple, blue, and green round rectangles, respectively. The network was constructed using Cytoscape tool. (b) Gene-gene interaction network of the genes associated with the zero-part SNPs and their interaction genes, as retrieved by BioGRID database. The genes associated with the zero-part SNPs and their interaction genes are depicted in round red and blue shapes, respectively, while the SNPs in purple rectangles. Larger node sizes indicate higher connectivity, while thicker edge sizes represent stronger evidence supporting the association. Drugs associated with the SNPs, according to PharmGKB variant annotations, are depicted in orange round rectangle shapes, and the chemical interactions of the genes, according to BioGRID database, are shown in green round shapes.

**Figure 4.**
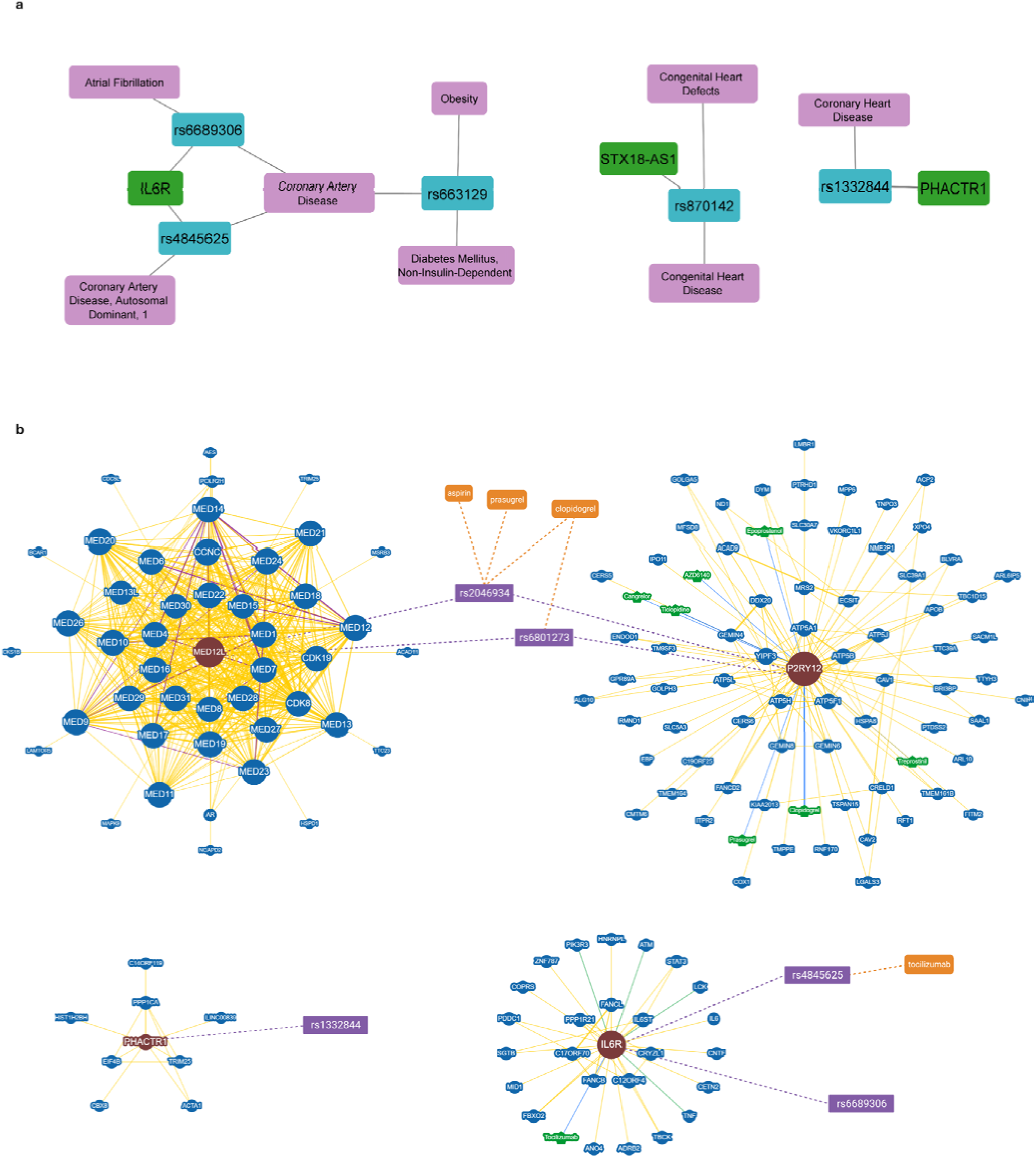
(a) Network visualization of the count-part SNPs and their association with diseases according to DisGeNET database. The disease annotations, the SNPs, and the genes corresponding to the SNPs are depicted in purple, blue, and green round rectangles, respectively. The network was constructed using Cytoscape tool. (b) Gene-gene interaction network of the genes associated with the count-part SNPs and their interaction genes, as retrieved by BioGRID database. The genes associated with the count-part SNPs and their interaction genes are depicted in round red and blue shapes, respectively, while the SNPs in purple rectangles. Larger node sizes indicate higher connectivity, while thicker edge sizes represent stronger evidence supporting the association. Drugs associated with the SNPs, according to PharmGKB variant annotations, are depicted in orange round rectangle shapes and the chemical interactions of the genes, according to BioGRID database, are shown in green round shapes.

The bioinformatic analysis of the genes associated to the zero-part model revealed the following associations: (i) the HDAC9 gene interacts with the non-selective histone deacetylase inhibitor panobinostat, and with anticonvulsive drug valproic acid (Fig. 3b); (ii) the Gene Ontology (GO) enrichment analysis based on Biological Process (BP) of the genes that interact with HDAC9 revealed that the interacting genes are significantly enriched in processes involved in protein deacetylation, histone modification, regulation of gene silencing by RNA and nuclear transport (Supplementary Fig. S2); (iii) a large number of genes that interact with *HFE* are mainly involved in glycoprotein biosynthetic and metabolic process, (Supplementary Fig. S3); (iv) the genes interacting with *ZPR1* are significantly enriched in biological processes involved in regulation of protein catabolic process and regulation of protein ubiquitination (Supplementary Fig. S4), whilst the variant rs964184 of *ZPR1* gene is associated with phenotypic modifications of the fenofibrate drug and antiretroviral agents via affecting *APOA1* and *APOA5* genes (Fig. 3b); (v) the genes that interact with *SMG6* are mainly enriched in biological processes related with RNA catabolic process, RNA localization/transport and regulation of translation (Supplementary Fig. S5); however, no drug interactions are recorded for the SMG6 rs216172 (Fig. 3b); (vi) the gene *CYP2C19* which encodes a cytochrome P450 enzyme involved in the metabolism of xenobiotics^30^ interacts with other genes of cytochrome P450 superfamily of enzymes (Fig. 3b) and; (vii) the SNP rs41291556 located in the *CYP2C19* gene affects the efficacy, toxicity and metabolism/pharmacokinetics of several drugs belonging to different pharmacologic classes including aspirin, clopidogrel, celecoxib, fluconazol etc. (Fig. 3b).

Concerning the count-part model, the significant associations extracted via the bioinformatics analysis are (i) the genes that interact with *MED12L* are significantly enriched in GO biological processes involved in the RNA biosynthetic processes and the initiation of DNA-dependent transcription initiation; the vast majority of the MED12L gene-interactors are subunit proteins of the Mediator complex which fundamental role is to communicate regulatory signals from DNA-bound transcription factors (TFs) to the RNA polymerase II (Pol II) enzyme^31^ (Supplementary Fig. S6). According to the PharmGKB database, the SNPs rs2046934 and rs6801273 mapped in the *MED12L* gene region are associated with phenotypic modifications of clopidogrel, aspirin, prasugrel (rs2046934), and clopidogrel (rs6801273), respectively (Fig. 4b); (ii) the genes that interact with *P2RY12* are involved mainly in oxidative phosphorylation biological process, and enrich pathways related to the respiratory electron transport, ATP synthesis by chemiosmotic coupling, and heat production by uncoupling proteins, as well as to the citric acid cycle and the mitochondrial biogenesis (Supplementary Fig. S7). Since the genes *MED12L* and *P2RY12* are OLGs in the human genome, the drug phenotype associated with the variants rs2046934 and rs6801273 are maintained the same as described above; (iii) the *IL6R* gene interacts with 25 genes, whilst rs4845625 affects the tocilizumab response of patients with rheumatoid arthritis^32^ (Fig. 4b) and; (iv) the *PHACTR1* gene interacts with eight genes, amongst them *HIST1, H2BH*, *PPP1CA*, *EIF4B*, and *TRIM25* (Fig. 4b).

The clinical features and the genetic characteristics found to be associated in the developed data-driven ML framework for the prediction and severity of CAD are depicted in Fig. 5.

**Figure 5.**
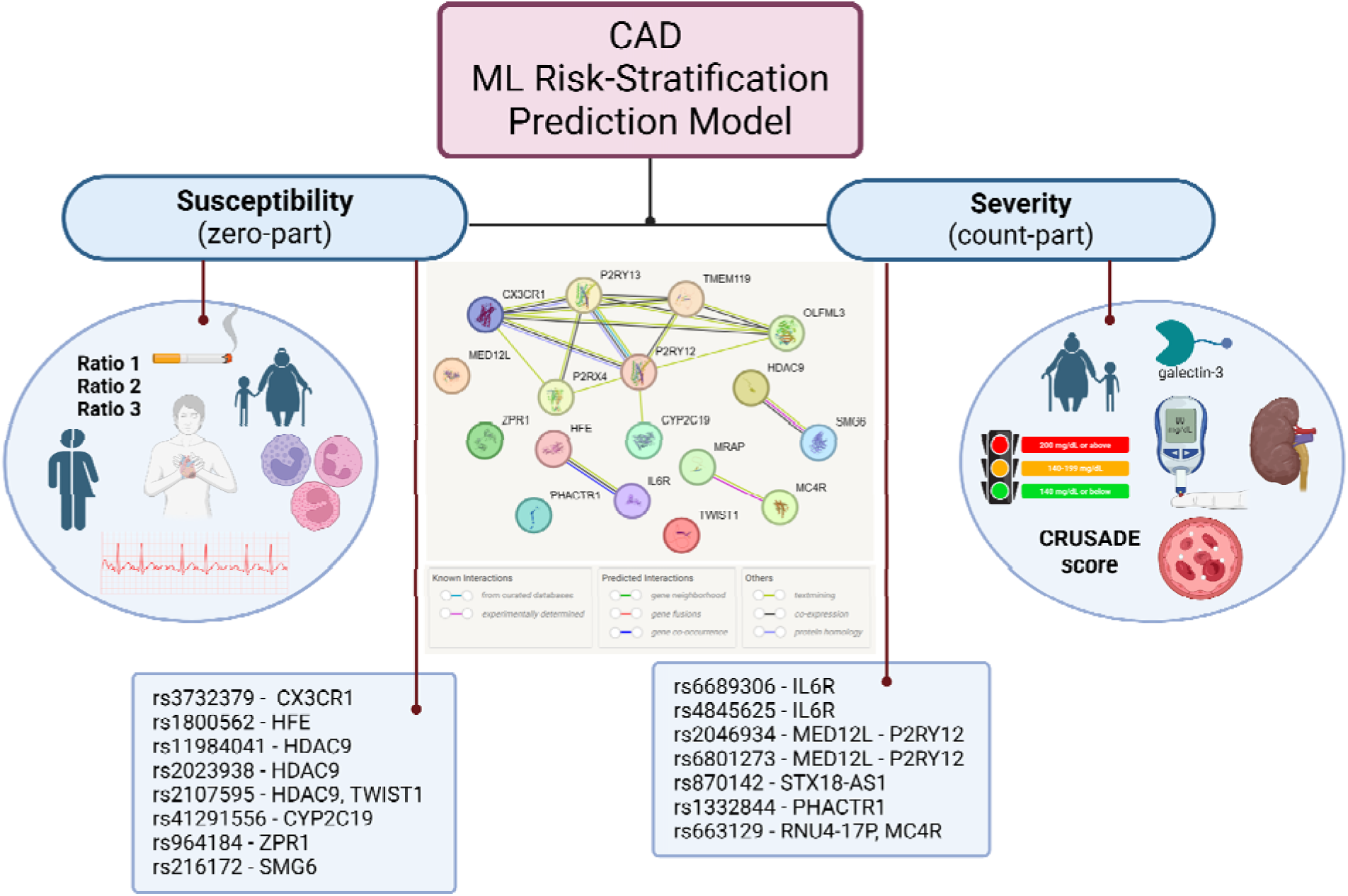
Schematic overview of the critical clinical, demographic and genetic features of the competing two-part ML risk-stratification model for the prediction of CAD. Note that the interaction between the genes of the identified important SNPs (shown in Table 1) were created through the protein-protein interaction networks functional enrichment analysis using the STRING database^77^.

## Discussion

The ML-based analysis presented herein is one of the first to demonstrate the enhanced predictive value of over 200 SNPs in addition to a clinically relevant predictive model for predicting angiographically confirmed obstructive CAD. Our results highlight that the combination of genetic information and clinical parameters, using a robust ML ensemble algorithm could effectively distinguish patients with and without obstructive CAD. As far as the genetic polymorphisms are concerned, the data obtained from the current study demonstrated for the first time eight out of 228 SNPs analyzed by NGS significantly associated with obstructive CAD occurrence and additional seven associated to the severity as expressed by higher SYNTAX score and, thus, increased CAD severity. To this end, the Boruta algorithm, implemented herein, yielded that adding these eight SNPs to a model consisting of fourteen clinical parameters could significantly bolster its predictive capability of obstructive CAD (zero-part model). Moreover, the addition of the second set of seven SNPs to a model consisting of nine clinically relevant risk-factors could also add to the predictive performance of the model for the prediction of the SYNTAX score (count-part model).

Regarding the zero-part model, the clinical variables associated with obstructive CAD were: patients’ age, gender, smoking history, atrial fibrillation history, the presence of chest pain or atypical angina symptoms, fasting glucose levels, neutrophil, eosinophil and basophil percentages, the ratio of monocyte to HDL-cholesterol (RATIO1), the ratio of lymphocyte to monocyte (RATIO2), the atherogenic index of plasma levels (RATIO3), and the ratio of SGOT to SGPT; (RATIO4). Of those variables, the presence of CAD-related symptoms (typical or atypical), higher patient’s age, gender, smoking history and higher lipidemic levels have been consistently used in most pre-test probability algorithms to serve as an effective gatekeeper for non-invasive testing and eliminate the likelihood of obstructive CAD without the need for invasive procedures^33,34^. The predictive significance of atrial fibrillation history has not been yet established; however, the overall incidence of obstructive CAD in patients presenting with atrial fibrillation is relatively high^35^. Additionally, higher fasting glucose levels could predict the occurrence of obstructive CAD in a chronic setting even if the observed hyperglycemia might be stress-induced^36,37^ in patients with suspected ACS. Moreover, the observed association between elevated levels of almost all subtypes of white blood cell counts (neutrophil, eosinophil and basophil percentages, and the ratio of lymphocyte to monocyte) and increased risk of obstructive CAD has been previously demonstrated as well^38,39^. This association might denote the underlying inflammation and/or hypersensitivity associated with CAD occurrence since leukocyte count is a widely available marker of inflammation.

As far as the count-part model of the Boruta algorithm is concerned, higher patients’ age, history of diabetes mellitus, elevated fasting glucose and galectin-3 levels, higher CRUSADE score, lower hemoglobin levels and chronic kidney disease (lower GFR, higher urea and creatinine values) were all associated with higher SYNTAX score. Patients’ age, diabetes mellitus history and dysglycemic status have been associated with increased CAD severity^40–42^. On the other hand, lower hemoglobin levels may have been already linked with poor prognostic course; however, no study has linked yet anemia (a potential inflammation marker) with CAD severity^43^. Furthermore, serum galectin-3 levels have been positively linked with CAD severity^44^, and renal dysfunction has been also associated with more severe CAD^45^ as demonstrated in our study. Finally, the positive association between CRUSADE and SYNTAX scores might be attributed to the already shown surrogate prognostic utility of both scores^46^.

Several ML-based predictive models have been already created aiming to predict CAD occurrence based on traditional or non-traditional CVD risk factors^47^. Promising approaches towards precision medicine currently investigate the predictive capacity of combined clinical and genomic data forced into ML-based algorithms. These investigations assessed genomic data (analysis of different SNPs) from real-world cohort studies^48–53^ or other registries such as U.K. BioBank^54,55^ and GWAS^56^. Although, most genetic risk score models are based on the genotyping of dozens of genetic loci, however, many of them do not show high specificity for the diagnosis of CAD^48^ and some of them do not add significant predictive utility on top of clinical CVD risk factors^53^.

As it was revealed from the bioinformatic and network analysis presented in this work, for the zero-part model, the significant eight SNPs, rs3732379, rs1800562, rs11984041, rs2023938, rs2107595, rs41291556, rs964184, and rs216172, have been previously associated with CAD, cerebrovascular events, hypertension, and diabetes mellitus^57^. Notably, three (rs11984041, rs2107595 and rs2023938) of the most important SNPs identified are associated with *HDAC9* gene that participates in deacetylation of histones, and, thus, regulates gene transcription^58^. Especially rs11984041 has been associated with large vessel occlusion in acute stroke^24^ and is in linkage disequilibrium with rs2107595. Both SNPs are found to elevate the risk for ischemic stroke via promoting carotid atherosclerosis^59^. However, a latter study in 733 large artery atherosclerotic stroke patients from China failed to confirm the significance of rs11984041^60^. In addition, the CARDIoGRAMplusC4D Consortium study identified 15 new susceptibility loci for CAD, amongst them rs2023938^61^. In a previous study of the same consortium rs964184 and rs216172 were denoted as significant susceptibility loci for CAD with risk allele frequencies of 0.13 and 0.37, respectively^62^. Of importance is also the rs1800562 in *HFE* gene, where a Mendelian randomization study highlighted its association between higher iron status and reduced CAD risk^63^.

Several GWASs performed between 2010 and 2015, as well as meta-analysis of their data, revealed several risk loci associated with CAD^22,27,61,64^. From our analysis, in the count-part model, the significant seven SNPs, rs6689306, rs4845625, rs2046934, rs6801273, rs870142, rs1332844, rs1332844 and rs663129, have been previously shown to be associated with CVDs, including CAD and atrial fibrillation. For instance, variants of the *IL6R* gene are associated with common diseases including inflammatory diseases such as CVDs^65^ and type 2 diabetes mellitus^66^. Specifically, rs6689306 is associated with atrial fibrillation, while rs4845625 is associated with the risk of CVDs including atrial fibrillation^67^, CAD^61^, and aortic aneurysm^65^. Two of the significant SNPs (rs2046934, rs6801273) are located at the overlapping genes *MED12L* and *P2RY12*. The latter is one of the most important pharmacological antiplatelet aggregation drug targets of crucial clinical value for CVD patients, where multiple SNPs, as well as different haplotypes, have been studied in terms of their significance with the risk of restenosis^68^, high on-treatment platelet reactivity^69^ and CAD^70^. The intergenic rs663129 is also associated with higher risk for CAD and diabetes mellitus, and with obesity-associated risk factors including lower HDL and higher triglyceride concentrations^71,72^. The rs870142 is involved in congenital heart diseases including ostium secundum atrial septal defect^27,73^. Finally, rs1332844, an intronic variant of the gene *PHACTR1* is also associated with CAD risk^74^ and major adverse cardiovascular events^75^.

In our study, the addition of SNPs to a clinical predictive model (Model A) offered statistically significant yet maybe not clinically relevant improvement in the diagnostic indices. This could be attributed to the fact that clinical CVD risk factors (i.e., dyslipidemia, diabetes mellitus, hypertension, obesity, and family history of CAD) *per se* represent the disease state and are often accompanied by multiple genetic correlations^56^. Dysregulation in any of those clinical parameters is probably a comprehensive manifestation of many genetic variations rather than the outcome of individual SNPs. Genetic variations eventually cause disease through CAD complexity (i.e., disturbed cellular balance, accumulation of harmful substances and damaged genetic cellular information). Hence, individuals with higher genetic risk scores are at higher risk of suffering from genotoxicity, which is not inevitable though therapeutic lifestyle change^76^, indicating that it must be the combined effect of various parameters determining the final phenotype, and CAD severity.

For the developed ML-risk stratification model to gain broad clinical applicability and practical utility, the external validation of our findings along with their clinical applicability in different patient populations must be demonstrated. To this end and in parallel to external validation, a definite feature analysis must be performed to allow practical scoring and interpretation of each predictor in a manner useful for clinicians in determining the risk of obstructive CAD. Also, future work must provide an explicit explanation for each predictor, thus, allowing the healthcare practitioner to read the result and decide according to the optimal cut point.

Despite the high-performance that our ML-risk stratification model shows in predicting the risk and severity of CAD by evaluating genetic, clinical, and demographic data, some study limitations warrant mention. Firstly, our data derived from a single center of similar ethnicity and the sample size was relatively small limiting the generalizability and universality of our findings. Secondly, this was a retrospective analysis of a prospectively designed study. Although the models could accurately classify individuals into obstructive and non-obstructive CAD, the predictive power of the models requires further validation in larger prospective studies.

## Conclusion

This study emphasizes that integrating clinical information with specific SNPs through ML can potentially assist in risk stratification for patients with suspected CAD. The selection of features required to build our ML-risk stratification model is likely to be the key determinant of its clinical performance. The addition of the genetic information to the clinical parameters further improves the performance of the model to predict the risk of CAD. Subsequently, the combination of such features can be used to build high-performance models for the prediction of CAD severity. Such an approach may enable healthcare professionals to identify patients not only at an increased risk for CAD but also for more complex forms of the disease, thereby facilitating targeted therapeutic interventions and preventive measures. In any case, however, this work also addresses the existing challenges in developing digitized precision medicine interventions related to the design of the trials, the data acquisition manner, as well as the clinical translation and implementation of molecular knowledge in healthcare. The development of such predictive models is envisaged to improve the accuracy and effectiveness of CVD prediction, ultimately leading to better patient outcomes broadly, positively affecting societal discrepancies and ultimately to a healthier population globally.

## Data Availability

All data produced in the present study are available upon reasonable request to the authors.

## Acknowledgements

This research has been co-financed by the European Regional Development Fund of the European Union and Greek national funds through the Operational Programme Competitiveness, Entrepreneurship, and Innovation, under the call RESEARCH–CREATE– INNOVATE (project code: T1EDK-02354).

## Authors’ contribution

Conceptualization of the study and ML model development: NM, LA, FC, ISV; Clinical study and data evaluation: GS, EK, ASP, NS; NGS analysis and data curation: FC, DC, ISV. Network and bioinformatics analysis: AD-G, AS, ISV; ML model validation and data interpretation: All authors contributed; Writing of the manuscript: All authors contributed; Supervision of the study: ISV; All authors have read and approved the final version of the manuscript.

## Note

Part of the data included in this article has been presented in an abstract in the “American Heart Association Scientific Sessions 2023” held in Philadelphia, PA, USA, 11-13 November 2023 (Chatzopoulou F, Mittas N, Karagiannidis E, Papazoglou AS, Stalikas N, Moysidis D, Giannopoulos-Dimitriou A, Saiti A, Ganopoulou M, Papa A, Chatzidimitriou D, Giannakoulas G, Sianos G, Angelis L, Vizirianakis IS. Abstract 17570: Combining genomic profiling and clinical data through machine learning modeling for the prediction of coronary artery disease severity: Insights from the GEnetic SYNTAX Score (GESS) trial. Circulation, 2023;148:A17570, published 6 Nov 2023); as well as in the 44^th^ Panhellenic Congress of Cardiology held in Thessaloniki, Greece, 12-14 October 2023.

## Supplementary Material

### List of Supplementary Tables

Supplementary Table 1. Selected clinical and demographic variables (features) from various types of Electronic Health Records (EHRs).

Supplementary Table 2. SNPs studied and their characteristics.

Supplementary Table 3. Descriptive statistics of elected clinical and demographic variables.

Supplementary Table 4. Results of statistical hypothesis testing procedures (first round of feature selection) for the subset of predictors with statistically significant effect on the distribution of SYNTAX score for the zero-part model.

Supplementary Table 5. Results of statistical hypothesis testing procedures (first round of feature selection) for the subset of predictors with statistically significant effect on the distribution of SYNTAX score for the count-part model.

Supplementary Table 6. Evaluation of prediction performances for competing models.

### List of Supplementary Figures

Supplementary Fig. S1. Distribution of SYNTAX score for the patients of the study (red) and for the patients with SYNTAX score higher than zero (blue).

Supplementary Fig. S2. Gene ontology (GO) and pathway enrichment analysis of the genes interacting with HDAC9 gene, according to BioGRID database.

Supplementary Fig. S3. Gene ontology (GO) and pathway enrichment analysis of the genes interacting with HFE gene, according to BioGRID database.

Supplementary Fig. S4. Gene ontology (GO) and pathway enrichment analysis of the genes interacting with ZPR1 gene, according to BioGRID database.

Supplementary Fig. S5. Gene ontology (GO) and pathway enrichment analysis of the genes interacting with SMG6 gene, according to BioGRID database.

Supplementary Fig. S6. Gene ontology (GO) and pathway enrichment analysis of the genes interacting with MED12L gene, according to BioGRID database.

Supplementary Fig. S7. Gene ontology (GO) and pathway enrichment analysis of the genes interacting with P2RY12 gene, according to BioGRID database.

### Supplementary Note

Machine Learning Framework

Bioinformatic and Network Analysis

