## Supplementary Tables 2-8 Figures 1-7 and Note for "Predicting coronary artery disease severity through genomic profiling and machine learning modelling: The GEnetic SYNTAX Score (GESS) trial"

**Running title:** ML prediction of genetic SYNTAX score

**Keywords:** Coronary artery disease, CAD, SYNTAX score, SNPs, biomarkers, cardiovascular disorders, CVDs, risk prediction, machine learning, personalized (precision) medicine, pharmacogenomics

-----------------------------------------------------------------------------------------------

* To whom correspondence should be addressed: Ioannis S. Vizirianakis, Laboratory of Pharmacology, School of Pharmacy, Aristotle University of Thessaloniki, Thessaloniki, Greece;; Tel.: 0030 2310 997658

Supplementary Fig. S2. Gene ontology (GO) and pathway enrichment analysis of the genes interacting with HDAC9, according to BioGRID database. (a) The top 15 significantly enriched gene ontology (GO) terms associated with the genes interacted with HDAC9. The gene ratio and statistical significance (p-value < 0.05, following Benjamini and Hochberg's adjustment method) are also depicted. (b) The REACTOME pathway enrichment analysis of the genes interacting with HDAC9. The top 15 statistically significant pathways are listed, and their colors correspond to the adjusted p-values. ........ 15

Supplementary Fig. S3. Gene ontology (GO) and pathway enrichment analysis of the genes interacting with HFE, according to BioGRID database. (a) The top 15 significantly enriched gene ontology (GO) terms associated with the genes interacting with HFE. The gene ratio and statistical significance (p-value < 0.05, following Benjamini and Hochberg's adjustment method) are also depicted. (b) The REACTOME pathway enrichment analysis of the genes interacting with HFE. The top 15 statistically significant pathways are listed, and their colors correspond to the adjusted p-values. ........ 16

Supplementary Fig. S4. Gene ontology (GO) and pathway enrichment analysis of the genes interacting with ZPR1, according to BioGRID database. (a) The top 15 significantly enriched gene ontology (GO) terms associated with the genes interacted with ZPR1. The gene ratio and statistical significance (p-value < 0.05, following Benjamini and Hochberg's adjustment method) are also depicted. (b) The REACTOME pathway enrichment analysis of the genes interacting with ZPR1. The top 15 statistically significant pathways are listed, and their colors correspond to the adjusted p-values. ........ 17

Supplementary Fig. S5. Gene ontology (GO) and pathway enrichment analysis of the genes interacting with SMG6, according to BioGRID database. (a) The top 15 significantly enriched gene ontology (GO) terms associated with the genes interacted with SMG6 gene. The gene ratio and statistical significance (p-value < 0.05, following Benjamini and Hochberg's adjustment method) are also depicted. (b) The REACTOME pathway enrichment analysis of the genes interacted with SMG6 gene. The top 15 statistically significant pathways are listed, and their colors correspond to the adjusted p-values. ... 18

Supplementary Fig. S6. Gene ontology (GO) and pathway enrichment analysis of the genes interacting with MED12L, according to BioGRID database. (a) The top 15 significantly enriched gene ontology (GO) terms associated with the genes interacted with MED12L. The gene ratio and statistical significance (p-value < 0.05, following Benjamini and Hochberg's adjustment method) are also depicted. (b) The REACTOME pathway enrichment analysis of the genes interacted with MED12L. The top 3 statistically significant pathways are listed, and their colors correspond to the adjusted p-values. ...................... 19

Supplementary Fig. S7. Gene ontology (GO) and pathway enrichment analysis of the genes interacting with P2RY12, according to BioGRID database. (a) Top 15 significantly enriched gene ontology (GO) terms associated with the genes interacted with P2RY12. The gene ratio and statistical significance (p-value < 0.05, following Benjamini and Hochberg's adjustment method) are also depicted. (b) The REACTOME pathway enrichment analysis of the genes interacting with P2RY12. The top 11 statistically significant pathways are listed, and their colors correspond to the adjusted p-values. ...................... 20

### Supplementary Note

Machine Learning Framework ........................................................................................................ 21

Bioinformatic and Network Analysis........................................................................................... 25

### Supplementary Tables

**Supplementary Table 1. Selected clinical and demographic variables (features) from various types of Electronic Health Records (EHRs)**

| EHR | Name | Description |
|---|---|---|
| HISTORY | GENDER | Gender |
| | HYPERTENSION | History of hypertension |
| | DIABETES MELLITUS | History of diabetes mellitus |
| | DYSLIPIDAEMIA | History of dyslipidaemia |
| | (+) FAMILY HISTORY | Positive (+) family history of CAD |
| | SMOKING | History of smoking |
| | AGE | Age of patient (in years) |
| | PREVIOUS STROKE | Previous stroke |
| | PERIPHERAL VASCULAR DISEASE | History of peripheral vascular disease |
| | AUTOIMMUNE DISEASE | History of any autoimmune disease |
| | ATRIAL FIBRILLATION | History of atrial fibrillation |
| ENTRY | CHEST PAIN | Chest pain |
| | ATYPICAL ANGINA SYMPTOMS | Atypical chest pain or shortness of breath |
| | BMI | Body mass index (kg/m$^2$) |
| | SAP | Systolic arterial pressure (SAP) (mmHg) |
| | DAP | Diastolic arterial pressure (DAP) (mmHg) |
| | CRUSADE SCORE | Crusade score |
| | QRS DURATION (ms) | QRS duration (in ms) |
| BIOCHEMICAL | GFR | Glomerular filtration rate by CKD-EPI (mL/min/1.73m2) |
| | GLU | Glucose (mg/dL) |
| | UREA | Urea (mg/dL) |
| | CREATININE | Creatinine (mg/dL) |
| | URIC ACID | Uric acid (mg/dL) |
| | CHOL | Total Cholesterol (mg/dL) |
| | LDL | Low density lipoprotein cholesterol (mg/dL) |
| | INR | International Normalized Ratio |
| | NEU% | Neutrophils percentage |
| | EOS% | Eosinophils percentage |
| | BASO% | Basophils percentage |
| | HGB | Hemoglobin (g/dL) |
| | MCV | Mean Corpuscular Volume (fL) |
| | MCH | Mean Corpuscular Hemoglobin (pg) |
| | RDW-CV | Red Blood Cell Distribution Width - coefficient of variation (percentage) |
| | RDW-SD | Red Blood Cell Distribution Width - standard deviation (percentage) |
| | PLT | Platelets (∗1000) |
| | MPV | Mean Platelet Volume (fL) |
| | PDW | Platelet Distribution Width (percentage) |
| | PCT | Plateletcrit (percentage) |
| | P-LCR | Platelet-large cell ratio |
| ESTIMATED RISK FACTORS | RATIO1 | Monocyte-to-HDL-cholesterol ratio |
| | RATIO2 | Lymphocyte-to-monocyte ratio |
| | RATIO3 | Atherogenic Index of Plasma levels, log(TG/HDL) |
| | RATIO4 | SGOT to SGPT ratio |

**Supplementary Table 2. SNPs studied and their characteristics**

Provided as separate attachment

**Supplementary Table 3. Descriptive statistics of elected clinical and demographic variables**

| | Number of patients with zero SYNTAX score (N=298) | Number of patients with non-zero SYNTAX score (N=655) | Total number of patients (N=953) |
|---|---|---|---|
| **SYNTAX score** | | | |
| N | 298 | 655 | 953 |
| Median (Q1, Q3) | - | 16.000 (9.000, 25.500) | 10.000 (0.000, 20.000) |
| **GENDER** | | | |
| N | 296 | 655 | 951 |
| Female | 111 (37.5%) | 141 (21.5%) | 252 (26.5%) |
| Male | 185 (62.5%) | 514 (78.5%) | 699 (73.5%) |
| Missing | 2 | 0 | 2 |
| **HYPERTENSION** | | | |
| N | 298 | 655 | 953 |
| No | 144 (48.3%) | 257 (39.2%) | 401 (42.1%) |
| Yes | 154 (51.7%) | 398 (60.8%) | 552 (57.9%) |
| **DIABETES MELLITUS** | | | |
| N | 298 | 654 | 952 |
| No | 240 (80.5%) | 463 (70.8%) | 703 (73.8%) |
| Yes | 58 (19.5%) | 191 (29.2%) | 249 (26.2%) |
| Missing | 0 | 1 | 1 |
| **DYSLIPIDAEMIA** | | | |
| N | 298 | 654 | 952 |
| No | 192 (64.4%) | 404 (61.8%) | 596 (62.6%) |
| Yes | 106 (35.6%) | 250 (38.2%) | 356 (37.4%) |
| Missing | 0 | 1 | 1 |
| **(+) FAMILY HISTORY** | | | |
| N | 297 | 654 | 951 |
| No | 251 (84.5%) | 532 (81.3%) | 783 (82.3%) |
| Yes | 46 (15.5%) | 122 (18.7%) | 168 (17.7%) |
| Missing | 1 | 1 | 2 |
| **SMOKING** | | | |
| N | 298 | 655 | 953 |
| No | 193 (64.8%) | 340 (51.9%) | 533 (55.9%) |
| Yes | 105 (35.2%) | 315 (48.1%) | 420 (44.1%) |
| **PREVIOUS STROKE** | | | |
| N | 298 | 654 | 952 |
| No | 291 (97.7%) | 633 (96.8%) | 924 (97.1%) |
| Yes | 7 (2.3%) | 21 (3.2%) | 28 (2.9%) |
| Missing | 0 | 1 | 1 |
| **AGE** | | | |
| N | 298 | 655 | 953 |

| | Number of patients with zero SYNTAX score (N=298) | Number of patients with non-zero SYNTAX score (N=655) | Total number of patients (N=953) |
|---|---|---|---|
| Median (Q1, Q3) | 64.000 (53.000, 72.000) | 65.000 (56.000, 73.000) | 65.000 (56.000, 73.000) |
| **PERIPHERAL VASCULAR DISEASE** | | | |
| N | 297 | 655 | 952 |
| No | 291 (98.0%) | 620 (94.7%) | 911 (95.7%) |
| Yes | 6 (2.0%) | 35 (5.3%) | 41 (4.3%) |
| Missing | 1 | 0 | 1 |
| **AUTOIMMUNE DISEASE** | | | |
| N | 298 | 655 | 953 |
| No | 292 (98.0%) | 645 (98.5%) | 937 (98.3%) |
| Yes | 6 (2.0%) | 10 (1.5%) | 16 (1.7%) |
| **ATRIAL FIBRILLATION** | | | |
| N | 298 | 655 | 953 |
| No | 248 (83.2%) | 597 (91.1%) | 845 (88.7%) |
| Yes | 50 (16.8%) | 58 (8.9%) | 108 (11.3%) |
| **BMI (kg/m2)** | | | |
| N | 298 | 653 | 951 |
| Median (Q1, Q3) | 27.800 (25.320, 31.185) | 28.100 (25.700, 30.900) | 28.000 (25.600, 31.000) |
| Missing | 0 | 2 | 2 |
| **SAP (mmHg)** | | | |
| N | 297 | 654 | 951 |
| Median (Q1, Q3) | 130.000 (120.000, 145.000) | 130.000 (120.000, 147.000) | 130.000 (120.000, 147.000) |
| Missing | 1 | 1 | 2 |
| **DAP (mmHg)** | | | |
| N | 297 | 654 | 951 |
| Median (Q1, Q3) | 80.000 (74.000, 87.000) | 79.000 (72.000, 85.750) | 80.000 (72.000, 86.500) |
| Missing | 1 | 1 | 2 |
| **CRUSADE SCORE** | | | |
| N | 292 | 647 | 939 |
| Median (Q1, Q3) | 20.000 (11.000, 33.000) | 21.000 (12.000, 33.000) | 21.000 (12.000, 33.000) |
| Missing | 6 | 8 | 14 |
| **QRS DURATION (ms)** | | | |
| N | 296 | 653 | 949 |
| Median (Q1, Q3) | 95.000 (88.000, 106.000) | 95.000 (87.000, 100.000) | 95.000 (88.000, 102.000) |
| Missing | 2 | 2 | 4 |
| **CHEST PAIN** | | | |
| N | 297 | 655 | 952 |
| No | 169 (56.9%) | 201 (30.7%) | 370 (38.9%) |

| | Number of patients with zero SYNTAX score (N=298) | Number of patients with non-zero SYNTAX score (N=655) | Total number of patients (N=953) |
|---|---|---|---|
| Yes | 128 (43.1%) | 454 (69.3%) | 582 (61.1%) |
| Missing | 1 | 0 | 1 |
| **ATYPICAL ANGINA SYMPTOMS** | | | |
| N | 297 | 655 | 952 |
| No | 193 (65.0%) | 500 (76.3%) | 693 (72.8%) |
| Yes | 104 (35.0%) | 155 (23.7%) | 259 (27.2%) |
| Missing | 1 | 0 | 1 |
| **GFR** | | | |
| N | 293 | 647 | 940 |
| Median (Q1, Q3) | 94.900 (76.700, 108.500) | 93.800 (70.500, 111.800) | 94.000 (72.800, 111.000) |
| Missing | 5 | 8 | 13 |
| **GLU (mg/dL)** | | | |
| N | 286 | 637 | 923 |
| Median (Q1, Q3) | 100.000 (90.250, 110.750) | 108.000 (94.000, 143.000) | 104.000 (93.000, 130.000) |
| Missing | 12 | 18 | 30 |
| **UREA (mg/dL)** | | | |
| N | 293 | 649 | 942 |
| Median (Q1, Q3) | 37.000 (30.000, 46.000) | 37.000 (29.000, 48.000) | 37.000 (29.000, 47.000) |
| Missing | 5 | 6 | 11 |
| **CREATININE (mg/dL)** | | | |
| N | 295 | 650 | 945 |
| Median (Q1, Q3) | 0.860 (0.750, 1.045) | 0.920 (0.780, 1.110) | 0.910 (0.770, 1.080) |
| Missing | 3 | 5 | 8 |
| **URIC ACID (mg/dL)** | | | |
| N | 173 | 340 | 513 |
| Median (Q1, Q3) | 5.800 (4.700, 6.800) | 5.800 (4.800, 6.925) | 5.800 (4.700, 6.900) |
| Missing | 125 | 315 | 440 |
| **CHOL (mg/dL)** | | | |
| N | 268 | 600 | 868 |
| Median (Q1, Q3) | 161.500 (140.000, 188.250) | 158.000 (132.000, 191.000) | 159.000 (134.000, 191.000) |
| Missing | 30 | 55 | 85 |
| **LDL (mg/dL)** | | | |
| N | 262 | 592 | 854 |
| Median (Q1, Q3) | 89.000 (70.000, 112.000) | 88.000 (64.000, 117.000) | 89.000 (66.000, 116.000) |
| Missing | 36 | 63 | 99 |
| **INR** | | | |
| N | 275 | 606 | 881 |
| Median (Q1, Q3) | 1.020 (0.970, 1.105) | 1.030 (0.980, 1.100) | 1.030 (0.980, 1.100) |

| | Number of patients with zero SYNTAX score (N=298) | Number of patients with non-zero SYNTAX score (N=655) | Total number of patients (N=953) |
|---|---|---|---|
| Missing | 23 | 49 | 72 |
| **NEU (%)** | | | |
| N | 294 | 651 | 945 |
| Median (Q1, Q3) | 61.950 (57.125, 66.750) | 66.000 (59.200, 72.700) | 64.200 (58.200, 71.100) |
| Missing | 4 | 4 | 8 |
| **EOS (%)** | | | |
| N | 294 | 650 | 944 |
| Median (Q1, Q3) | 2.100 (1.025, 2.900) | 1.550 (0.700, 2.700) | 1.700 (0.800, 2.800) |
| Missing | 4 | 5 | 9 |
| **BASO (%)** | | | |
| N | 294 | 650 | 944 |
| Median (Q1, Q3) | 0.400 (0.200, 0.600) | 0.300 (0.200, 0.500) | 0.400 (0.200, 0.500) |
| Missing | 4 | 5 | 9 |
| **HGB (g/dL)** | | | |
| N | 294 | 651 | 945 |
| Median (Q1, Q3) | 13.900 (12.700, 14.875) | 13.700 (12.600, 14.900) | 13.800 (12.700, 14.900) |
| Missing | 4 | 4 | 8 |
| **MCV (μm$^3$)** | | | |
| N | 294 | 651 | 945 |
| Median (Q1, Q3) | 87.400 (84.200, 89.975) | 87.200 (83.600, 90.200) | 87.300 (83.800, 90.100) |
| Missing | 4 | 4 | 8 |
| **MCH (pg per cell)** | | | |
| N | 294 | 651 | 945 |
| Median (Q1, Q3) | 29.550 (28.300, 30.600) | 29.500 (28.400, 30.700) | 29.500 (28.400, 30.700) |
| Missing | 4 | 4 | 8 |
| **RDW-CV (%)** | | | |
| N | 290 | 639 | 929 |
| Median (Q1, Q3) | 13.400 (12.900, 14.300) | 13.500 (12.900, 14.400) | 13.500 (12.900, 14.400) |
| Missing | 8 | 16 | 24 |
| **RDW-SD (fL)** | | | |
| N | 294 | 649 | 943 |
| Median (Q1, Q3) | 43.100 (40.525, 45.000) | 42.700 (40.400, 45.400) | 42.800 (40.400, 45.300) |
| Missing | 4 | 6 | 10 |
| **PLT (× 10$^9$/L)** | | | |
| N | 293 | 651 | 944 |
| Median (Q1, Q3) | 219.000 (189.000, 267.000) | 231.000 (190.000, 275.000) | 227.500 (189.750, 273.000) |
| Missing | 5 | 4 | 9 |
| **MPV (fL)** | | | |
| N | 286 | 636 | 922 |

| | Number of patients with zero SYNTAX score (N=298) | Number of patients with non-zero SYNTAX score (N=655) | Total number of patients (N=953) |
|---|---|---|---|
| Median (Q1, Q3) | 10.900 (10.325, 11.700) | 10.800 (10.200, 11.500) | 10.800 (10.200, 11.600) |
| Missing | 12 | 19 | 31 |
| **PDW (%)** | | | |
| N | 286 | 636 | 922 |
| Median (Q1, Q3) | 13.000 (12.000, 15.000) | 13.000 (12.000, 15.000) | 13.000 (12.000, 15.000) |
| Missing | 12 | 19 | 31 |
| **PCT (%)** | | | |
| N | 286 | 635 | 921 |
| Median (Q1, Q3) | 0.240 (0.210, 0.290) | 0.250 (0.210, 0.290) | 0.250 (0.210, 0.290) |
| Missing | 12 | 20 | 32 |
| **P-LCR (%)** | | | |
| N | 280 | 616 | 896 |
| Median (Q1, Q3) | 32.800 (28.175, 38.800) | 32.400 (27.000, 37.800) | 32.500 (27.400, 38.125) |
| Missing | 18 | 39 | 57 |
| **RATIO1** | | | |
| N | 265 | 597 | 862 |
| Median (Q1, Q3) | 1.283 (0.946, 1.767) | 1.765 (1.256, 2.438) | 1.580 (1.133, 2.270) |
| Missing | 33 | 58 | 91 |
| **RATIO2** | | | |
| N | 294 | 650 | 944 |
| Median (Q1, Q3) | 0.309 (0.240, 0.393) | 0.351 (0.253, 0.493) | 0.334 (0.249, 0.462) |
| Missing | 4 | 5 | 9 |
| **RATIO3** | | | |
| N | 266 | 600 | 866 |
| Median (Q1, Q3) | 0.382 (0.218, 0.572) | 0.520 (0.352, 0.712) | 0.484 (0.312, 0.665) |
| Missing | 32 | 55 | 87 |
| **RATIO4** | | | |
| N | 289 | 641 | 930 |
| Median (Q1, Q3) | 1.042 (0.773, 1.359) | 1.148 (0.875, 1.687) | 1.125 (0.846, 1.556) |
| Missing | 9 | 14 | 23 |
| **GALECTIN (ng/mL)** | | | |
| N | 298 | 655 | 953 |
| Median (Q1, Q3) | 17.350 (13.600, 22.550) | 18.000 (14.100, 23.300) | 17.900 (13.900, 23.200) |
| Values represent number of patients (N), number of missing observations if any (Missing), median and interquartile range (Median (interquartile range)) for continuous characteristics and number of patients (percentage) for categorical characteristics | | | |

**Supplementary Table 4. Results of statistical hypothesis testing procedures (first round of feature selection) for the subset of predictors with statistically significant effect on the distribution of SYNTAX score for the zero-part model**

| **Input Type** | **Predictor** | **Method** | ***Statistic*** | ***p*** |
|---|---|---|---|---|
| SNP | rs2023938 | LR | 14.843 | 0.001 |
| | rs2107595 | LR | 12.904 | 0.002 |
| | rs41291556 | LR | 9.044 | 0.003 |
| | rs11984041 | LR | 10.122 | 0.006 |
| | rs1330344 | LR | 7.641 | 0.022 |
| | rs25882 | LR | 7.320 | 0.026 |
| | rs10455872 | LR | 4.725 | 0.030 |
| | rs964184 | LR | 6.586 | 0.037 |
| | rs1042522 | LR | 6.548 | 0.038 |
| | rs4252185 | LR | 6.290 | 0.043 |
| | rs3732379 | LR | 6.262 | 0.044 |
| | rs445925 | LR | 6.150 | 0.046 |
| | rs216172 | LR | 6.066 | 0.048 |
| | rs1042713 | LR | 5.535 | 0.063 |
| | rs17228212 | LR | 5.448 | 0.066 |
| | rs1800562 | LR | 3.217 | 0.073 |
| | rs2900478 | LR | 5.202 | 0.074 |
| | rs4516035 | LR | 5.070 | 0.079 |
| | rs4363657 | LR | 4.946 | 0.084 |
| | rs116843064 | LR | 4.934 | 0.085 |
| | rs361525 | LR | 4.864 | 0.088 |
| | rs2306374 | LR | 4.638 | 0.098 |
| Clinical | GENDER | LR | 25.786 | <0.001 |
| | SMOKING | LR | 13.902 | <0.001 |
| | CHEST PAIN | LR | 58.381 | <0.001 |
| | ATYPICAL ANGINA SYMPTOMS | LR | 12.956 | <0.001 |
| | GLU | LR | 46.332 | <0.001 |
| | NEU (%) | LR | 25.108 | <0.001 |
| | RATIO1 (Monocyte-to-HDL-cholesterol ratio) | LR | 54.375 | <0.001 |
| | RATIO2 (Lymphocyte-to-monocyte ratio) | LR | 19.919 | <0.001 |
| | RATIO3 (Atherogenic Index of Plasma) | LR | 43.601 | <0.001 |
| | RATIO 4 (SGOT to SGPT ratio) | LR | 41.650 | <0.001 |
| | DIABETES MELLITUS | LR | 10.434 | 0.001 |
| | ATRIAL FIBRILLATION | LR | 12.090 | 0.001 |
| | HYPERTENSION | LR | 6.901 | 0.009 |
| | BASO (%) | LR | 6.656 | 0.010 |
| | PERIPHERAL VASCULAR DISEASE | LR | 6.250 | 0.012 |
| | AGE | LR | 3.360 | 0.067 |
| | EOS (%) | LR | 3.084 | 0.079 |
| | HGB | LR | 3.041 | 0.081 |
| | MPV | LR | 3.001 | 0.083 |

**Supplementary Table 5. Results of statistical hypothesis testing procedures (first round of feature selection) for the subset of predictors with statistically significant effect on the distribution of SYNTAX score for the count-part model**

| Input Type | Predictor | Method | *Statistic* | *p* |
|---|---|---|---|---|
| SNP | rs1801133 | K-W | 10.053 | 0.007 |
| | rs2948080 | K-W | 8.427 | 0.015 |
| | rs663129 | K-W | 8.459 | 0.015 |
| | rs3184504 | K-W | 8.170 | 0.017 |
| | rs4845625 | K-W | 7.815 | 0.020 |
| | rs3798220 | M-W | 8601 | 0.020 |
| | rs6689306 | K-W | 7.552 | 0.023 |
| | rs1803274 | K-W | 7.070 | 0.029 |
| | rs2046934 | K-W | 6.761 | 0.034 |
| | rs10495809 | K-W | 6.692 | 0.035 |
| | rs6725887 | K-W | 6.636 | 0.036 |
| | rs870142 | K-W | 6.326 | 0.042 |
| | rs12190287 | K-W | 5.578 | 0.061 |
| | rs1057910 | K-W | 5.556 | 0.062 |
| | rs6801273 | K-W | 5.165 | 0.076 |
| | rs1332844 | K-W | 4.991 | 0.082 |
| | rs1800849 | K-W | 4.986 | 0.083 |
| | rs6903956 | K-W | 4.901 | 0.086 |
| | rs1122608 | K-W | 4.785 | 0.091 |
| | rs1412444 | K-W | 4.673 | 0.097 |
| | rs56062135 | K-W | 4.663 | 0.097 |
| Clinical | DIABETES MELLITUS | M-W | 35211.5 | 0.000 |
| | CHEST PAIN | M-W | 40945 | 0.036 |
| | PERIPHERAL VASCULAR DISEASE | M-W | 8899 | 0.073 |
| | AGE | Spearman | 0.174 | <0.001 |
| | CRUSADE SCORE | Spearman | 0.178 | <0.001 |
| | GFR | Spearman | -0.160 | <0.001 |
| | GLU | Spearman | 0.177 | <0.001 |
| | UREA | Spearman | 0.158 | <0.001 |
| | GALECTIN-3 | Spearman | 0.136 | <0.001 |
| | CREATININE | Spearman | 0.103 | 0.009 |
| | HGB | Spearman | -0.086 | 0.029 |
| | NEU (%) | Spearman | 0.068 | 0.084 |

**Supplementary Table 6. Evaluation of prediction performances for competing models**

| | | Metric | | | | | |
|---|---|---|---|---|---|---|---|
| **Base learner** | **Predictors** | **Accuracy** | **Precision** | **Recall** | **$F_1$** | **$AUC_{ROC}$** | **$AUC_{PR}$** |
| Zero-part (existence of obstructive CAD) | Clinical (Model A) | 72.4% | 76.9% | 88.3% | 82.2% | 0.757 | 0.893 |
| | Clinical+SNPs (Model B) | 78.2% | 80.2% | 92.6% | 85.9% | 0.798 | 0.912 |
| | | **Metric** | | | | | |
| **Base learner** | **Predictors** | MdAE | | | MdMRE | | |
| Count-part (severity of CAD) | Clinical (Model A) | 7.00 | | | 52.19% | | |
| | Clinical+SNPs (Model B) | 6.96 | | | 49.99% | | |
| **Ensemble** | **Predictors** | MdAE | | | | | |
| Overall | Clinical (Model A) | 7.61 | | | | | |
| | Clinical+SNPs (Model B) | 7.00 | | | | | |

### Supplementary Figures

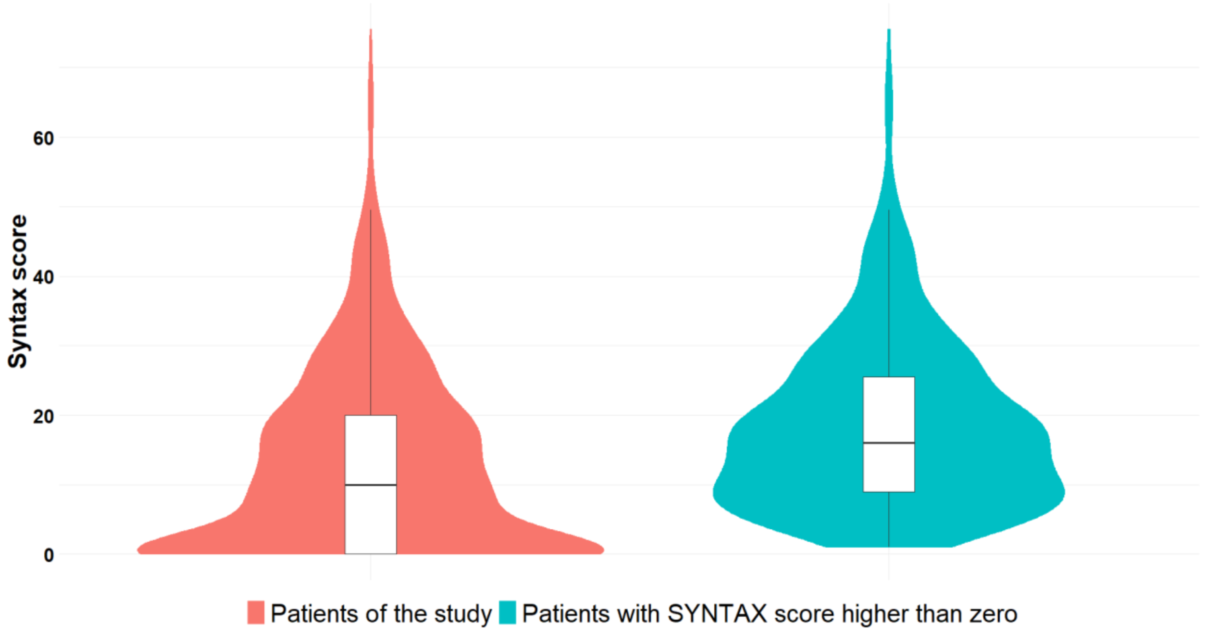

**Supplementary Fig. S1.** Distribution of SYNTAX score for the patients of the study (red) and for the patients with SYNTAX score higher than zero (blue).

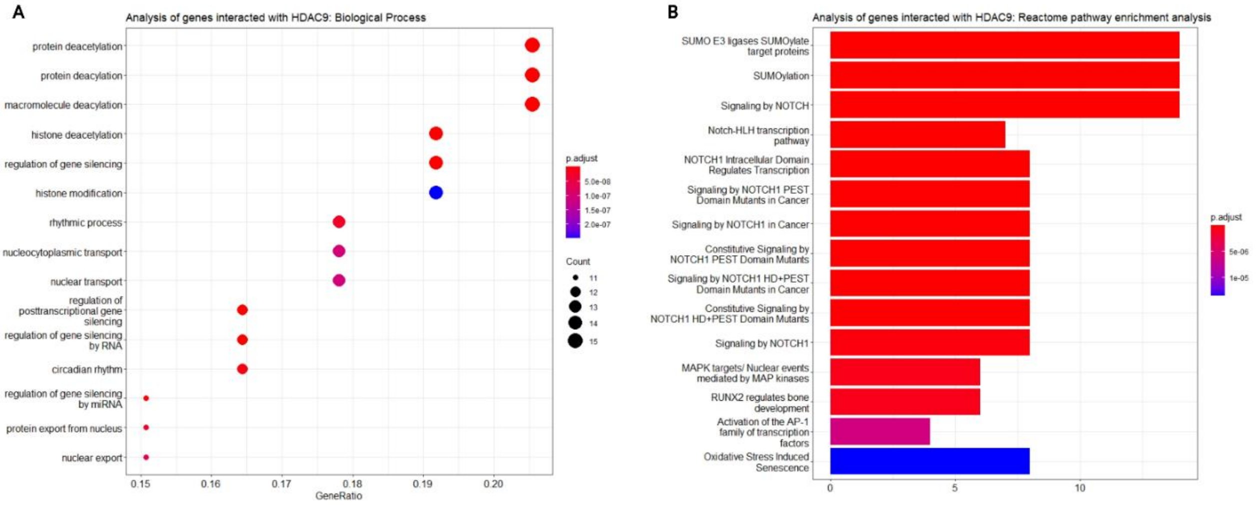

**Supplementary Fig. S2. Gene ontology (GO) and pathway enrichment analysis of the genes interacting with *HDAC9* gene, according to BioGRID database. (a)** Top 15 significantly enriched gene ontology (GO) terms associated with the genes interacted with *HDAC9*. The gene ratio and statistical significance (p-value < 0.05, following Benjamini and Hochberg's adjustment method) are also depicted. **(b)** The REACTOME pathway enrichment analysis on the genes interacted with *HDAC9*. The top 15 statistically significant pathways are listed, and their colors correspond to the adjusted p-values.

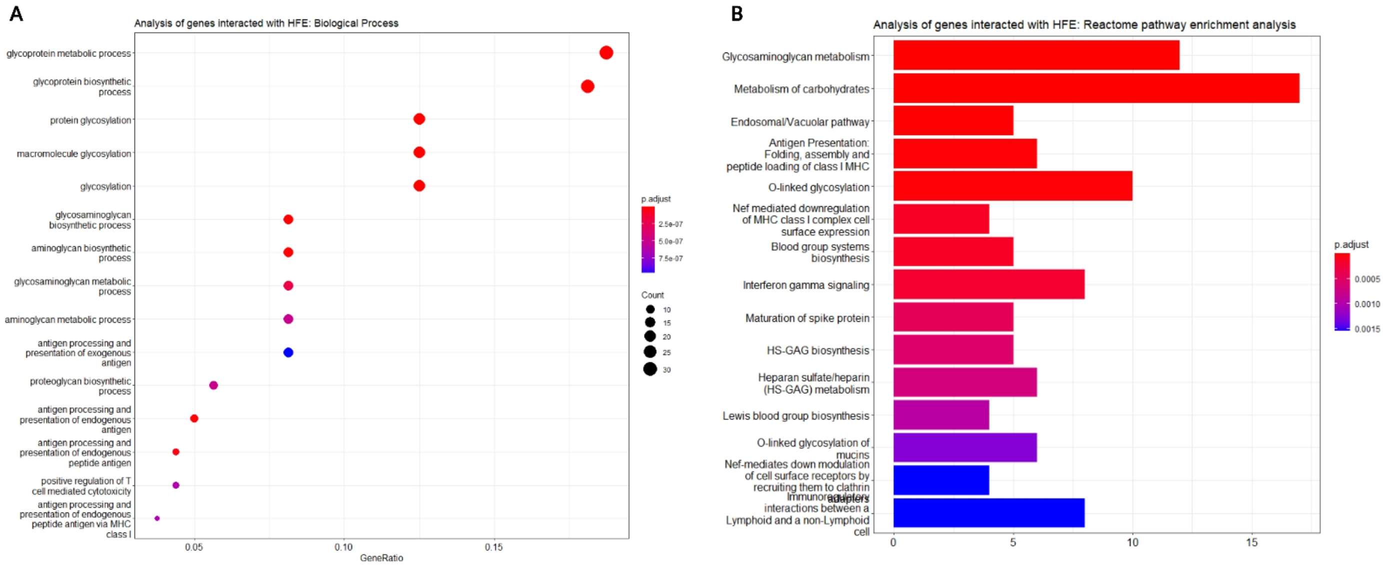

**Supplementary Fig. S3. Gene ontology (GO) and pathway enrichment analysis of the genes interacting with *HFE* gene, according to BioGRID database. (a)** Top 15 significantly enriched gene ontology (GO) terms associated with the genes interacting with *HFE*. The gene ratio and statistical significance (p-value < 0.05, following Benjamini and Hochberg's adjustment method) are also depicted. **(b)** The REACTOME pathway enrichment analysis on the genes interacting with *HFE*. The top 15 statistically significant pathways are listed, and their colors correspond to the adjusted p-values.

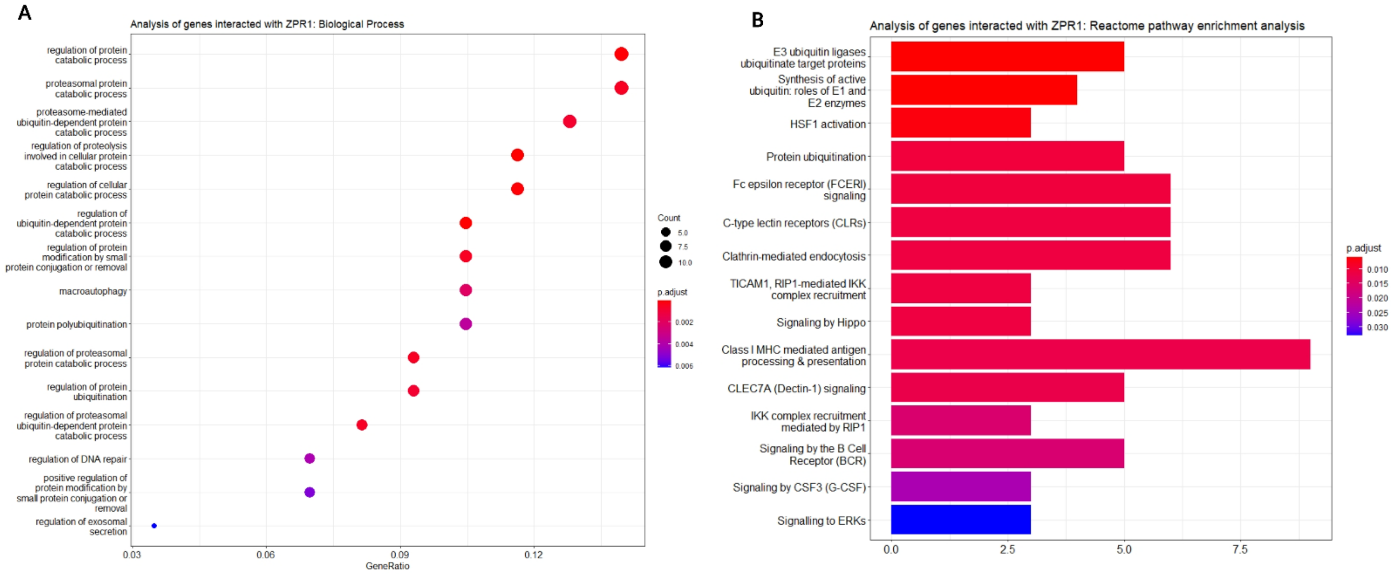

**Supplementary Fig. S4. Gene ontology (GO) and pathway enrichment analysis of the genes interacting with *ZPR1* gene, according to BioGRID database. (a)** Top 15 significantly enriched gene ontology (GO) terms associated with the genes interacting with *ZPR1*. The gene ratio and statistical significance (p-value < 0.05, following Benjamini and Hochberg's adjustment method) are also depicted. **(b)** The REACTOME pathway enrichment analysis on the genes interacted with *ZPR1*. The top 15 statistically significant pathways are listed, and their colors correspond to the adjusted p-values.

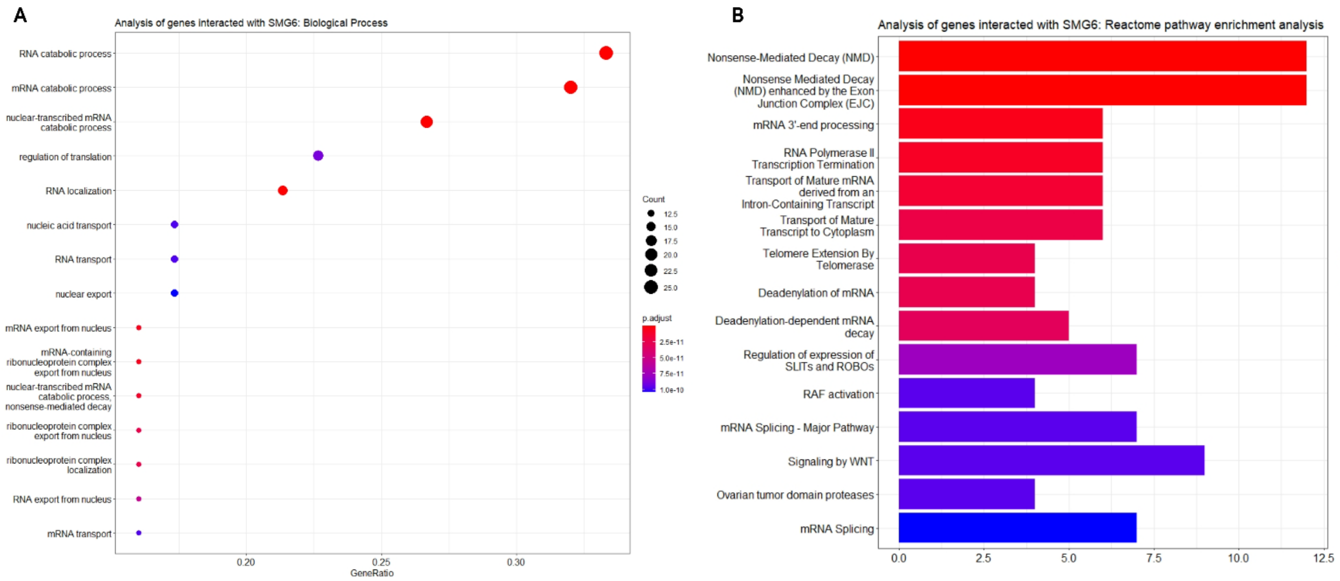

**Supplementary Fig. S5. Gene ontology (GO) and pathway enrichment analysis of the genes interacting with *SMG6* gene, according to BioGRID database. (a)** The top 15 significantly enriched gene ontology (GO) terms associated with the genes interacting with *SMG6*. The gene ratio and statistical significance (p-value < 0.05, following Benjamini and Hochberg's adjustment method) are also depicted. **(b)** The REACTOME pathway enrichment analysis on the genes interacting with the *SMG6*. The top 15 statistically significant pathways are listed, and their colors correspond to the adjusted p-values.

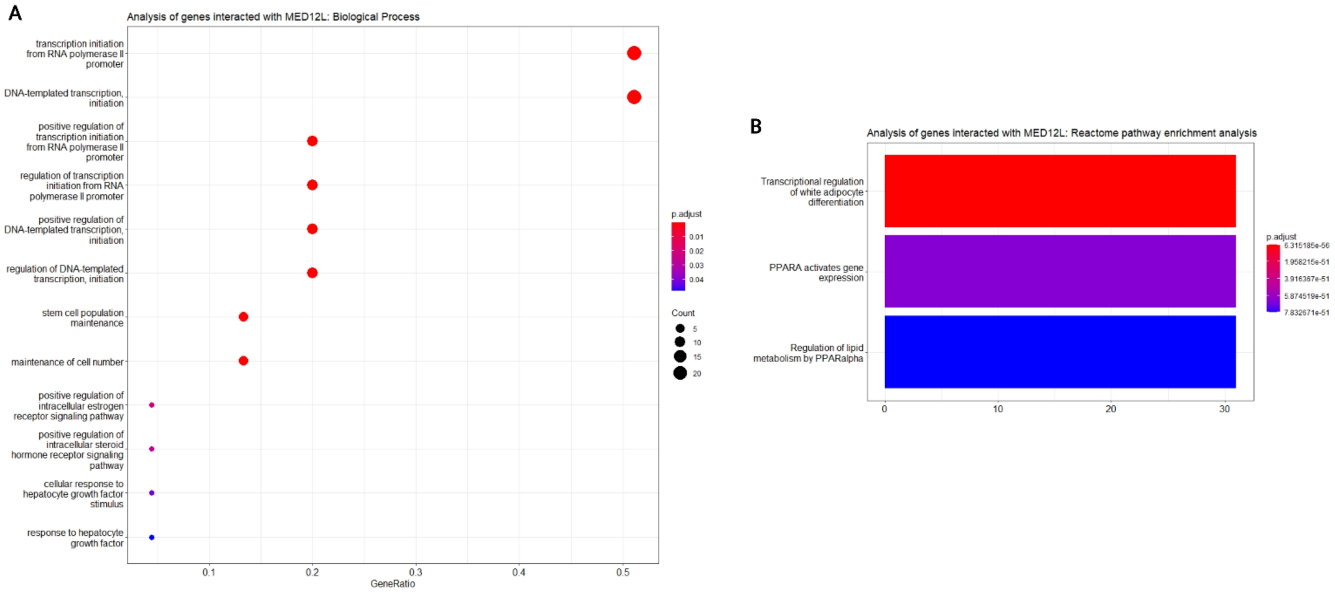

**Supplementary Fig. S6. Gene ontology (GO) and pathway enrichment analysis of the genes interacting with *MED12L* gene, according to BioGRID database. (a)** The top 15 significantly enriched gene ontology (GO) terms associated with the genes interacting with *MED12L*. The gene ratio and statistical significance (p-value $< 0.05$, following Benjamini and Hochberg's adjustment method) are also depicted. **(b)** The REACTOME pathway enrichment analysis of the genes interacting with *MED12L*. The top 3 statistically significant pathways are listed, and their colors correspond to the adjusted p-values.

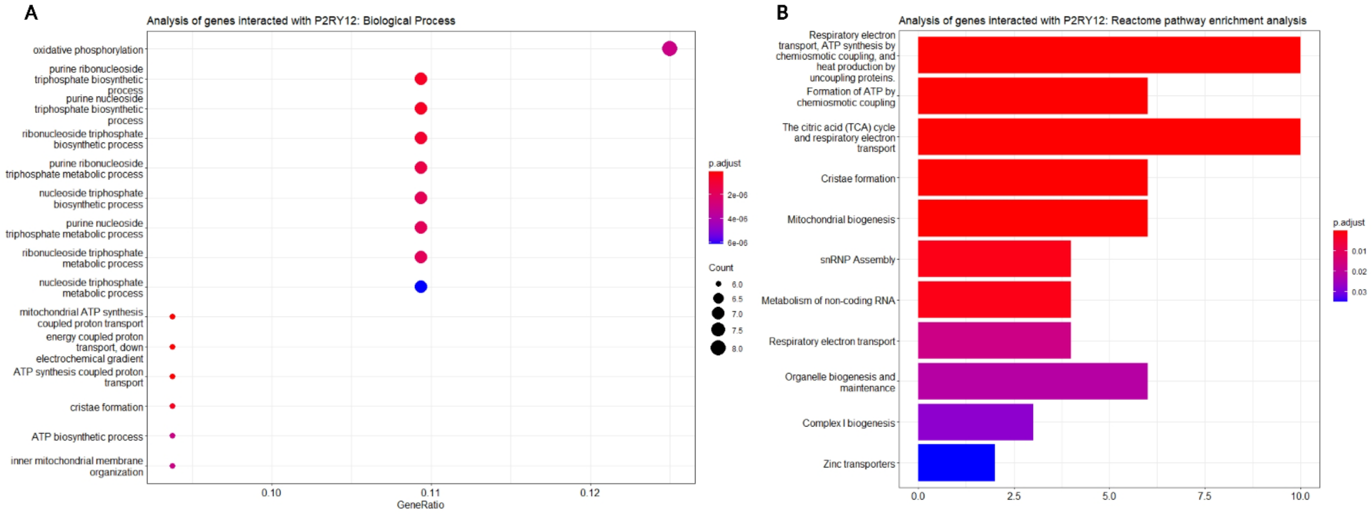

**Supplementary Fig. S7. Gene ontology (GO) and pathway enrichment analysis of the genes interacting with *P2RY12* gene, according to BioGRID database. (a)** The top 15 significantly enriched gene ontology (GO) terms associated with the genes interacted with *P2RY12*. The gene ratio and statistical significance (p-value < 0.05, following Benjamini and Hochberg's adjustment method) are also depicted. **(b)** The REACTOME pathway enrichment analysis on the genes interacted with *P2RY12*. The top 11 statistically significant pathways are listed, and their colors correspond to the adjusted p-values.

### Supplementary Note

#### Machine Learning Framework

This section is dedicated to presenting the phases of the ML framework adopted for integrating clinical and genetic biomarkers, with the aim of leveraging the wealth of *Electronic Health Records* (EHRs) (Supplementary Table 1) and information hidden in genomic signatures derived from SNPs (Supplementary Table 2) in the Data Collection phase. During the Data Preparation phase, various pre-processing steps were carried out, such as labeling, cleaning, removal, or correction of erroneous entries etc. to ensure the quality of the resulting ML solution.

The next phase, Data Exploration, involved the application of descriptive and exploratory analytics (Supplementary Table 3) for gaining insights into the collected variables (or features) that characterize different aspects of the examined population. The primary focus was to investigate the SYNTAX score and its empirical distribution to draw conclusions about its key characteristics including central tendency, variability and shape. The graphical inspection (Supplementary Fig. 1) revealed a right-skewed distribution with 31.27% ($N = 298$) of patients exhibiting a SYNTAX score of zero. The relatively high proportion of zero values in the SYNTAX score distribution, indicating a zero-inflated response variable, was the primary reason for our choice not to pursue a straightforward ML solution. Instead, we sought an alternative approach to address the challenges associated with the excess of zero values. Due to the above issues, a generic ensemble approach (Model Selection) was proposed [1] separating the building phase into two distinct processes (or models): the zero-part model, which is responsible for classifying patients into two groups (zero vs. non-zero SYNTAX score) based on the absence or presence of obstructive CAD and the count-part model, which is dedicated to evaluating of the severity of CAD, as indicated by the SYNTAX score, for patients classified in the non-zero SYNTAX score group. This means that the count-part model was fitted only on training instances (i.e., patients) from the truncated-at-zero SYNTAX score distribution. In real-world applications, the prediction of SYNTAX score for a new patient is generated via the synergy of the two described above. First, the zero-part model is applied to predict whether the patient should be classified as a non-zero or zero case. If the patient is predicted to have a non-zero SYNTAX score, the count-part model, then, provides an estimated value for the severity of SYNTAX score.

Besides the main advantage of the proposed approach in handling the zero-inflated SYNTAX score distribution, the proposed approach also offers flexibility in identifying and evaluating potentially diverse sets of risk factors that may influence (a) the absence or presence of obstructive CAD and (b) the severity of CAD. In fact, feature selection is considered one of the most impactful tasks in the ML lifecycle, as the massive amount of information hidden in clinical EHRs and genomic data, expressed through identified SNPs, presents challenges for building an effective solution. On the one hand, a high number of features can be proved beneficial for uncovering undiscovered patterns of significant practical importance for decision-makers and healthcare providers. On the other hand, the curse of multidimensionality may degrade the prediction capabilities of a ML model, leading to poor generalization, while the inclusion of redundant features may increase the complexity of the model.

To identify important risk factors (predictors) for inclusion into the Model Building phase, a two-step feature selection protocol was designed (Feature Selection) utilizing both traditional statistical methods (Step 1) and advanced feature selection mechanisms (Step 2). In the first step, appropriate hypothesis testing procedures were applied to identify features that did not present a statistically significant effect on the distribution of SYNTAX score, thus failing to meet the inclusion criteria for the second round of the feature selection process. This step accounted for the types of response variables and the set of predictors involved in the two-part modeling approach. For the case of the zero-part model, the identification of risk factors presenting a significant effect on the response variable was accomplished via separate univariate logistic regression models for the total set of the candidate features (Supplementary Table 4). In contrast, for the case of the count-part model, non-parametric hypothesis testing procedures were applied, due to the fact that SYNTAX score presented a non-normal and positively skewed truncated-at-zero distribution. Specifically, the *Spearman's rho correlation coefficient* was used for examining the relationships between the response and the set of continuous predictors, whereas the non-parametric *Mann-Whitney U* test and *Kruskal-Wallis* test were employed for identifying factors (i.e., categorical features) with two and more than two levels, respectively, that presented a significant effect on the SYNTAX score distribution. In this first step, we applied a less strict alpha level criterion that was set at 0.10.

The qualified sets of predictors from both the zero-part and count-part models were, further, subjected to a second selection round to determine the final set of predictors to be inserted into the Model Building phase. To achieve this objective, the Boruta algorithm [2], a feature selection method belonging to the general branch of wrapper methods, was employed separately for the zero- and count-part models, as described in our previous study [1]. The reasons to employ the Boruta method rather than other feature selection mechanisms are discussed below. First, the approach can be used for feature selection purposes in both classification and regression tasks, which is a key requirement, in our context, as the proposed approach integrates both types of prediction tasks into a unified model. Second, it is a modification of the well-known *Random Forest* (RF) algorithm that is able to capture not only linear relationships but also complex non-linear associations between the response and the set of predictors, as well as interaction effects. Third, it is based on an iterative data-generation process that produces, at each repetition, a new version of the original dataset augmented with randomly shuffled (or permuted) values of all features (*shadow features*). After fitting a RF model to the merged dataset, which combines both the original and the shadow features, the algorithm evaluates the *variable importance measure* (VIM) of each original feature against the performance accomplished by the "*best*" shadow feature. Lastly, the algorithm results into a set of important (or informative) features based on well-established statistical concepts such as the binomial distribution and traditional hypothesis testing procedures, handling with an efficient manner the uncertainty that is inherent in the decision-making process. In summary, the second step of the feature selection protocol can be viewed as a refinement further mechanism for identifying the important predictors that should be considered when building the ensemble model.

Regarding the next phase (Model Building), there is a wide range of ML approaches that can serve as candidate algorithms for fitting the zero- and count-parts models. The following criteria were predefined in our selection process: (a) the algorithm should be applicable for both classification (zero-part model) and regression (count-part model) tasks; (b) the algorithm should be capable of capturing both linear and non-linear relationships between the predictors and the response variable, (c) the algorithm should effectively cope with mixed-type data (d) the algorithm should have been successfully deployed in other application domains. In addition to these requirements, extensive experimentation with the data suggested that *eXtreme Gradient Boosting* (XGBoost) [3], which is a well-known ML framework that optimizes the stochastic gradient boosting algorithm [4] through an iterative approach, generally provided superior predictive capabilities compared to other ML algorithms. The tuning phase of the algorithm for both the zero- and the count-part models was based on a grid search strategy, while a 3-fold cross-validation approach was adopted for the selection of the models' hyper-parameters.

Considering the mixed nature of the proposed approach, the prediction capabilities of the two competing models (without/with SNPs), on the test set, were evaluated using appropriate performance metrics commonly used in classification and regression tasks. For the zero-part models, four performance indicators (Accuracy, Precision, Recall and F1-score) derived from the confusion matrix were assessed. Additionally, *Receiver Operating Characteristic* (ROC) [5] and *Precision-Recall* (PR) curves [6] were constructed to facilitate a graphical investigation of the achieved performances. The predictive power of the count-part models was evaluated using performance metrics that quantify the deviance between the actual $y_i$ and predicted $\hat{y}_i$ values for each test case in the dataset. More specifically, two loss functions (absolute error-AE and magnitude of relative error-MRE) were considered to compute an overall performance indicator by aggregating the derived errors for each test case using the central tendency measure of the median. To evaluate the prediction capabilities of the examined models, we employed a *hold-out* schema using 2/3 of the cases for fitting each model (training phase) and the remaining 1/3 of data for evaluating performance on unseen cases (testing phase).

Finally, since the focal research question of the study was whether practitioners should incorporate genomic information, in addition to clinical data, into existing data-driven solutions to enhance decision-making processes, we conducted appropriate statistical hypothesis testing procedures in order to infer about the generalizability of the derived findings. The competing models were assessed for their discriminating power using the *Area Under the Curve* (AUC) via *Delong's* test, while the error distributions derived from the count-part models were compared through the non-parametric *Wilcoxon Signed Rank* test.

After having succeed to evaluate the predictive capacity of the two competing models, the important SNP variants identified via the zero-part and count-part models were subjected into bioinformatic analysis by retrieving the implicated genes from the ClinVar public archive[1].

[1] https://www.ncbi.nlm.nih.gov/clinvar/ accessed on 20 April 2023

The human diseases associated with the important predicted SNPs were recovered via the DisGeNET platform[2] (v7.0) utilizing the disgenet2r package (v099.2) in R. Only the data derived from expert-curated databases integrated within DisGeNET database were retrieved and appropriately filtered to include only disease entries mapped to the following UMLS® (Unified Medical Language System®) semantic types: Disease or Syndrome, Neoplastic Process, Acquired Abnormality, Anatomical Abnormality, Congenital Abnormality and Mental or Behavioral Dysfunction. The Cytoscape (v3.9.0) software platform was used for disease-variant network construction and visualization. The open-access database BioGrid[3] containing expert-curated experimental evidence from focused low-throughput and large high-throughput interactome studies was used to construct networks depicting the genetic, protein and chemical interactions of the genes affected by the important SNPs. In addition, curated variant-drug pairs were recovered from PharmGKB database[4] according to variant annotations (association between a variant and a drug phenotype) and integrated in the interactome networks. *Gene Ontology* (GO) and pathway enrichment analysis was performed for the interaction genes of the genes corresponding to the SNPs utilizing the ClusterProfiler and ReactomePA packages, respectively.

**Bioinformatic and Network Analysis**

The Gene Ontology (GO) and pathway enrichment analysis, according to BioGRID[2] database of the genes associated with the identified important SNPs of the ML risk-stratification model for the prediction of susceptibility and severity of CAD, is shown in Supplementary Figures S2-S7 and their data is presented within the main text of the manuscript. The bioinformatic analysis of the SNPs corresponding to zero-part model, according to DisGeNET database, revealed that most of the SNPs are associated with CVDs. Importantly, rs2107595 is associated with peripheral arterial disease, CAD, hypertension, cerebrovascular events and Moyamoya disease, a progressive steno-occlusive intracranial disorder caused by blocked arteries in the basal ganglia area, which can lead to CAD [7]. Ischemic stroke and cerebrovascular events have been also associated with rs11984041, while rs2023938 is related only with CAD. Indeed, *Genome-Wide Association Studies* (GWAS) identified the gene

---

[2] https://www.disgenet.org/ accessed on 20 April 2023
[3] https://thebiogrid.org/ accessed on 20 April 2023
[4] https://www.pharmgkb.org/ accessed on 20 April 2023

region of HDAC9 as a major risk locus and the rs2107595 as the leading SNP associated with stroke and CAD [8,9]. Interestingly, in vivo experiments have reported that rs2107595 has allele-specific transcriptional capacity and is related to enhanced expression of HDAC9 via the allele-specific recruitment of the E2F3/Rb1/TFDP1 complex to the rs2107595 region. This mechanism is explained by a physical interaction of the rs2107595 region with the HDAC9 gene promoter region, promoting the expression of the HDAC9 gene in an allele-specific manner [10]. Regarding the SNPs of HDAC9 gene, rs11984041 has been linked with large-vessel ischemic stroke in individuals with European ancestry, according to a GWAS [11].

The analysis of the SNP, mapped to SMG6 gene region, unveiled an involvement of rs216172 to CAD. The most extended SNP-disease networks were created by rs964184 and rs1800562, suggesting that these SNPs affects the normal functioning of many biological pathways. Particularly, rs964184 is involved in metabolic syndrome, diabetes mellitus, hyperlipidemia, and malignant neoplasms. Concerning CVDs, rs964184 is associated with CAD and hypertension. Moreover, rs964184 has been strongly associated with serum lipid concentrations in fasting and postprandial state as well as with increased MI risk. However, a recent study suggested that a low-fat diet can modulate the effect of genetic variability in the rs964184 of ZPR1 gene on the altered postprandial triglyceride response [12]. Although many diseases are associated with rs1800562, including neoplastic disease, porphyria, iron binding capacity functions, hemochromatosis, Alzheimer and Diabetes mellitus, the only relation of this SNP with cardiovascular risk is through its association with elevated diastolic blood pressure [13].

The protein and genetic interactions of the genes affected by the important SNPs predictors of the zero- and count-part components were retrieved from BioGRID expert-curated database and integrated into graphical network representations (Fig. 3 and Fig. 4). The GO analysis based on *Biological Process* (BP) of the genes that interact with HDAC9 revealed that the interacting genes are significantly enriched in processes involved in protein diacylation, histone modification, regulation of gene silencing by RNA and nuclear transport (Supplementary Fig. S2). The pathway analysis revealed that the interacted genes of the HDAC9 proteins participate in pathways mainly involved in post-translational protein modification, signal transduction (signaling by NOTCH, etc.) and MAP kinase activation. Based on the chemical interaction data retrieved from BioGRID the HDAC9 gene interacts with the non-selective histone deacetylase inhibitor Panobinostat as well as with anticonvulsive drug valproic acid (Fig. 3b). The genes interacting with the ZPR1 gene are significantly enriched in biological processes involved in regulation of protein catabolic process, regulation of protein ubiquitination etc., as well as in pathways associated with Protein ubiquitination, signal transduction (signaling by Hippo, etc.), innate immune system (Fc epsilon receptor signaling, Class I MHC mediated antigen processing & presentation etc.) (Supplementary Fig. S4). According to PharmGKB database, the SNP rs964184 identified in the ZPR1 is associated with phenotypic modifications of the fenofibrate drug and antiretroviral agents via affecting APOA1 and APOA5 genes located within the APOA1/C3/A4/A5-ZPR1-BUD13 gene cluster (Fig. 3b). The genes interacting with HFE gene are significantly enriched in biological processes involved in glycoprotein biosynthetic and metabolic process, aminoglycan biosynthetic and metabolic process etc., as well in pathways related with glycosaminoglycan metabolism, metabolism of carbohydrates, endosomal/vacuolar pathway etc (Supplementary Fig. S3). According to PharmGKB database, the SNP rs1800562 is associated with phenotypic modifications of the mineral iron and stimulating glycoprotein epoetin alfa (Fig. 3b). The genes interacted with SMG6 are mainly enriched in biological processes related with RNA catabolic process, RNA localization/transport, regulation of translation etc., as well as in pathways involved in Nonsense-Mediated Decay, transport of mature transcript, mRNA processing/splicing etc (Supplementary Fig. S5). In terms of the drugs associated with the SNP rs216172 located in SMG6 gene, no drug interactions are recorded in PharmGKB database. The gene CYP2C19 which encodes a cytochrome P450 enzyme involved in the metabolism of xenobiotics interacts with other genes of cytochrome P450 superfamily of enzymes. The SNP rs41291556 located in the CYP2C19 gene affects the efficacy, toxicity, and metabolism/PK of several drugs belonging to different pharmacologic classes including aspirin, clopidogrel, celecoxib, fluconazol etc (Fig. 3b). The CX3CR1 gene encoding the fractalkine receptor displays a compact interaction network with the genes HIF1A, CCDC167 and CX3CL1 (Fig. 3b).
